# Proteomic prediction of disease largely reflects environmental risk exposure

**DOI:** 10.1101/2025.08.27.25334571

**Authors:** Kristin Tsuo, M. Austin Argentieri, Danni Gadd, Mitja Kurki, Zhili Zheng, Denis Baird, Riccardo E. Marioni, Christopher Foley, Hailiang Huang, Benjamin B. Sun, Chia-Yen Chen, Mark J. Daly, Alicia R. Martin

## Abstract

Plasma proteomic signatures accurately predict disease risk, but our understanding of the mechanisms contributing to the predictive value of the proteome remains limited. Here, we characterized proteomic biomarkers of 19 age-related diseases, based on observational associations between 2,923 protein levels and incidence of these outcomes in the UK Biobank (N = 45,438). To identify the subset of these biomarkers that may represent causal drivers of disease, we first employed Mendelian Randomization (MR) and found that only 8% of the protein-disease associations with genetic instruments showed suggestive evidence of causal relationships, and were more likely to pertain to only a single disease. We then tested the hypothesis that many proteomic biomarkers, particularly the non-causal proteins, are impacted by environmental factors that might independently affect disease risk and protein levels. We discovered that the vast majority (>90%) of proteins associated with diseases like lung cancer and COPD are also associated with smoking, and more than half of all disease-associated proteins tested in MR were associated with smoking. These proteins showed no evidence of causal effects on disease, suggesting their predictive value is as an environmental sensor. Given the sensitivity of the plasma proteome to smoking, we developed a proteomic score for smoking (SmokingPS) and demonstrated that the plasma proteome can serve as a quantitative index of smoking behavior and history. Extending this approach to alcohol intake phenotypes, our results generally suggest that many plasma proteins identified in observational associations are more likely to be readouts of environmental risk factors than disease-specific signals. We conclude that the plasma proteome may provide critical objective biomarkers for quantifying the impacts of environmental risk factors on human health and disease. Our results have significant implications for implementing predictive plasma protein biomarkers in disease prevention, and can help guide interpretation of putative protein-disease associations as actionable therapeutic targets or quantitative indications of upstream exposures that represent potential intervention points.

## Introduction

Technological advances in high-throughput, broad-capture proteomic assays have highlighted the potential of leveraging plasma proteins for disease prediction, biomarker discovery, and therapeutic development. In the past few years, population-scale biobanks have used these assays to profile portions of the plasma proteome in thousands of individuals^1–5^. The UK Biobank Pharma Proteomics Project (UKB-PPP) generated one of the largest proteomic resources to date, with measurements of nearly 3,000 blood plasma analytes in more than 54,000 UKB participants using the Olink Explore 3072 proximity extension assay^6^. Coupled with existing deep phenotyping data, UKB-PPP creates a unique opportunity to interrogate the links between the plasma proteome and a broad spectrum of diseases and disease-related traits. Indeed, studies have already leveraged this resource to identify thousands of associations between plasma protein levels and disease outcomes^7–9^. Proteomic predictors of incident disease, developed from these associations, outperform traditional clinical models and polygenic risk scores for a wide range of diseases^7,8,10^ (e.g. AUC > 0.8 for 92 diseases^9^).

The ability of the plasma proteome to predict incident disease is promising for a range of clinical applications – for example building on ELISA (enzyme-linked immunosorbent assay) and other multiplex immunoassays for diagnostics and monitoring treatments^11^, as well as for patient stratification in clinical trials^12^ – but our understanding of what contributes to the predictive value of the circulating proteome remains limited. A few overlapping mechanisms could explain the widespread associations between plasma proteins and incident disease outcomes: (1) plasma proteins have direct causal effects on disease onset; (2) plasma proteins are impacted by disease processes that begin before disease diagnosis; and (3) plasma proteins are impacted by external factors, like environmental exposures, that both contribute to disease onset and affect protein levels. Gaining a deeper understanding of the specific roles of proteomic biomarkers in disease prediction has important implications for the potential clinical utility of these biomarkers.

First, plasma proteins with causal effects on disease may be candidates for novel therapeutic targets. Several disease-specific studies have investigated the potential causal roles of plasma proteins, including for type II diabetes^13^, cardiometabolic diseases^10,14,15^, psychiatric disorders^16,17^, and gastrointestinal diseases^18,19^. Phenome-wide studies across many diseases and phenotypes have also investigated the causal basis of protein-disease associations^20,21^. These studies have identified putative drug targets such as SCARA5 for cardioembolic stroke^15^ and IL1RL1 for inflammatory bowel diseases^20^.

On the other hand, non-causal plasma proteins may serve as biomarkers for monitoring disease onset and progression. For example, a study on plasma proteomic biomarkers of dementia found that abundances of certain predictive proteins began changing years before diagnosis^22^. Non-causal plasma proteins may also potentially help monitor impacts of changes in lifestyle and other environmental factors on disease risk. Previous work has speculated that the plasma proteome captures environmental effects^7,23^, and a handful of studies investigating factors contributing to the variance of individual proteins have found that environmental exposures can explain a substantial portion of variability in the levels of certain proteins^24,25^. However, major gaps remain in understanding how modifiable factors contribute to the utility of the overall proteome as disease predictors. In particular, few studies to date have simultaneously evaluated putatively causal proteins alongside other mechanisms that may explain the predictive value of non-causal associations. In this study, we aimed to conduct a comprehensive characterization of the relationships between plasma proteins and key disease outcomes in order to delineate proteins as potential therapeutic targets or non-causal biomarkers of environmental risk factors.

Utilizing blood proteomic data available from a subset of the UKB-PPP cohort (N = 45,438, see **Methods**), we investigated an expanded set of associations between 2,923 unique proteins and 23 age-related incident disease outcomes (see study design in **Figure 1**). We first applied Mendelian Randomization (MR) to partition the disease-associated proteomic biomarkers into those with potential causal roles across diseases (i.e. drivers) vs. non-causal roles (i.e. predictors). Next, we examined the impacts of smoking on the plasma proteome because of its large environmental effect on disease risk. Integrating the MR and smoking analyses, we identified that a large proportion of protein predictors are exposure-associated and likely play no causal role in disease pathogenesis. To further evaluate the sensitivity of the plasma proteome to smoking, we developed a proteomic score for smoking, and tested this score alongside smoking status and other clinical biomarkers in disease prediction models. By replicating this proteomic score in an external dataset (FinnGen), we highlight the objective, quantitative, and generalizable nature of environmental risk quantification enabled by proteomics. Finally, we confirmed the generality of this paradigm by developing and testing an additional proteomic score for alcohol intake, which is nearly entirely independent from the smoking protein score.

**Figure 1.**
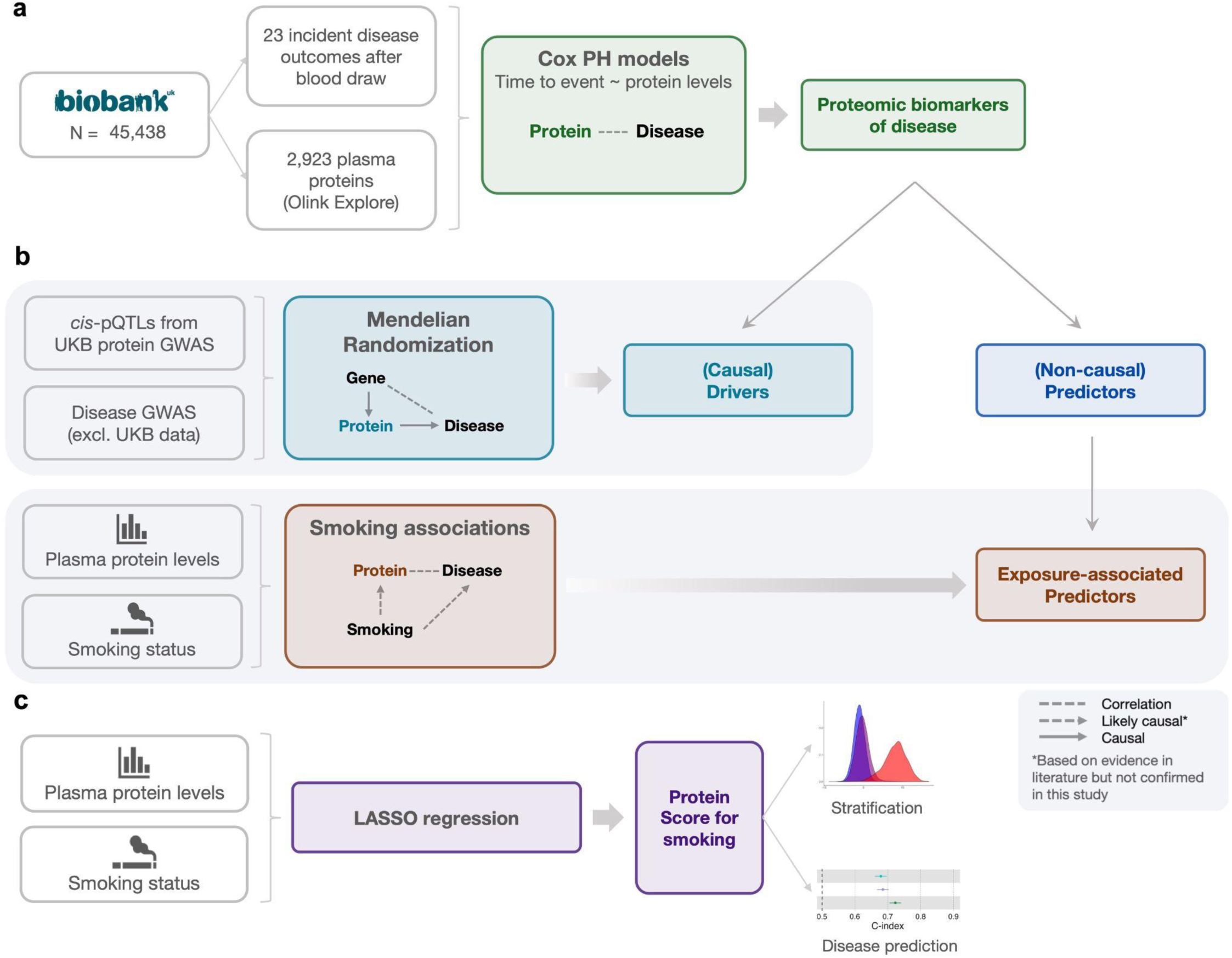
Study design for characterizing proteomic biomarkers of disease. **a,** We tested for associations between 2,923 plasma proteins measured in a subset of UKB-PPP participants and 23 incident disease outcomes using Cox proportional hazards (PH) models. We refer to the proteins in the significant protein-disease associations as biomarkers. **b,** To better understand the protein-disease associations identified in **(a)**, we took two approaches. First, we identified which of the protein-disease associations were likely causal by applying two-sample Mendelian Randomization (MR) using protein quantitative trait loci (pQTLs) as genetic instruments and disease GWAS that did not utilize UKB data; we refer to the putatively causal proteins as drivers. Second, we identified proteins that are likely not causal themselves but instead reflect the effects of disease-related exposures, by identifying proteins that were significantly associated with smoking but lacking evidence from MR; we refer to these proteins as exposure-associated predictors. **c,** We trained a LASSO regression model on the subset of UKB-PPP participants with smoking status data to develop a protein score for smoking (SmokingPS). We demonstrated that the SmokingPS accurately captures quantity and frequency of smoking and used SmokingPS to predict disease incidence.

## Results

### Plasma proteins are widely associated with incident disease

Previously, we tested associations between levels of 1,468 plasma proteins, measured in UKB-PPP participants, and 23 incident diseases over a 15-year follow-up period using Cox proportional hazards (PH) models. In this study, we expanded these analyses to include associations with an additional 1,455 proteins (total proteins = 2,923), measured in the same individuals. Details on the plasma protein measurements, QC procedures, and UKB-PPP sample have been described previously^6^ (see also **Methods**). We found 9,308 significant associations between 2,122 proteins and 22 of the 23 incident disease outcomes, adjusting for age and sex variables (Bonferroni-adjusted *P-*value < 0.05/(23*2923) = 7.44 x 10^-7^) (**Supplementary Table 1**). The number of associations ranged from 1 association each for ALS, multiple sclerosis, colorectal cancer, and major depressive disorder to 1,653 and 1,798 for liver disease and type 2 diabetes, respectively; brain/CNS cancer showed no significant protein associations (**Extended Data** Fig. 1).

### A small subset of proteins have causal effects on disease

To identify which subset of the thousands of protein-disease associations discovered represent potentially causal relationships, we applied two-sample Mendelian Randomization (MR). 19 of the 23 disease outcomes that we tested in the Cox PH models had suitable GWAS summary statistics for MR, as they did not include UKB data which was used to identify genetic instruments (**Supplementary Table 2**). These diseases are the focal set for subsequent analyses. We leveraged the catalog of protein quantitative trait loci (pQTLs) that were previously mapped to these proteins^6^ to first subset to proteins with robust genetic instruments. To minimize potential biases, we restricted genetic instruments to *cis-*pQTLs, and retained the proteins with *F*-statistics > 10 and that passed Steiger filtering. Ultimately, 2,907 of the significant protein-disease pairs (involving 782 unique proteins) had genetic instruments and were tested for causality via MR (**Supplementary Table 3**).

Only 225/2,907 protein-disease pairs (8%) showed suggestive evidence for causality (*P*-value < 0.05) (**Figure 2a**). In additional sensitivity tests for horizontal pleiotropy using MR-Egger^26^, we found that 9 of the 225 nominally significant protein-disease pairs may have pleiotropic genetic instruments (**Supplementary Table 4**), and thus further validation of these pairs may be needed. Overall, these *cis-*MR analyses did not reveal causal roles for the majority of the proteins identified in observational associations, indicating that the vast majority of protein-disease associations likely cannot be explained by causal effects of the protein on disease onset.

**Figure 2.**
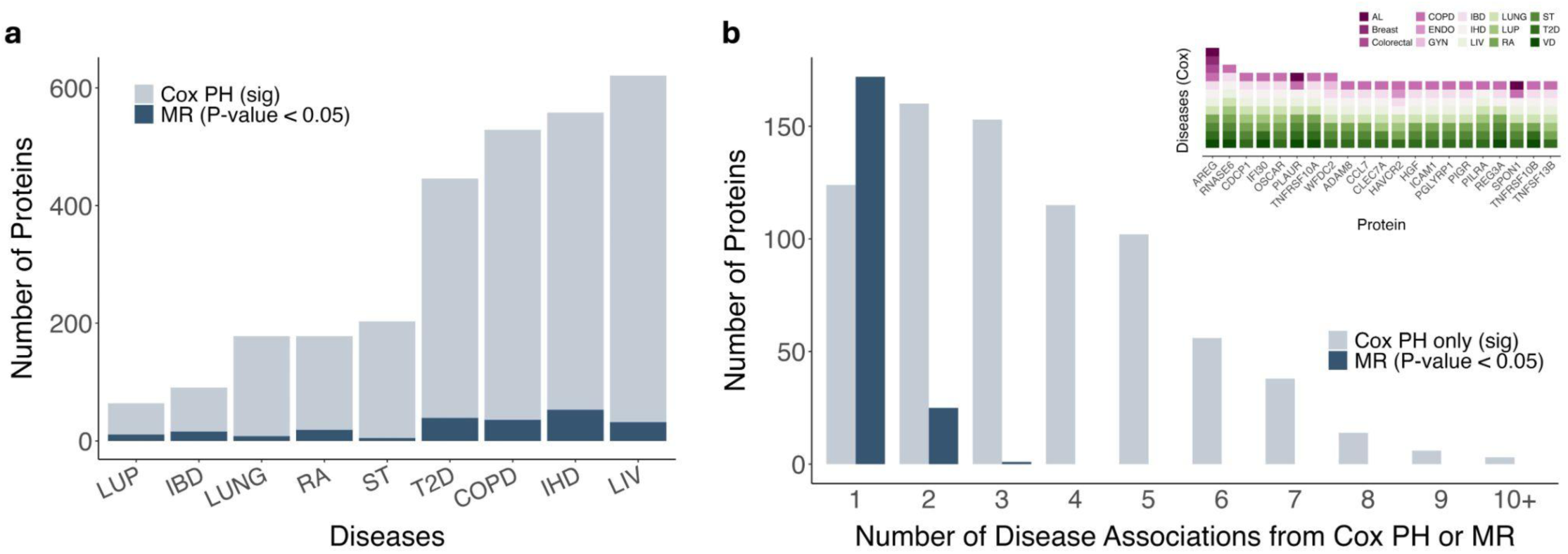
Comparison of proteins involved in observational vs. causal associations across diseases. **a,** Counts of proteins significantly associated with diseases in Cox PH models and that also had genetic instruments (N = 782 proteins involved in 2,907 protein-disease pairs). Diseases with more than 20 significant protein associations are shown. Counts of the subset of these proteins showing suggestive evidence of causality from MR analyses (*P*-value < 0.05) are indicated by darker blue bars. **b,** Counts of proteins associated with 1 to more than 10 diseases in the Cox PH models vs. MR tests. All proteins shown here are involved in the 2,907 protein-disease pairs that were significant in Cox PH models and had genetic instruments. Light blue bars represent non-causal predictors; dark blue bars represent causal drivers. Inset shows 27 proteins significantly associated with 8 or more diseases in the Cox PH models (IBD = inflammatory bowel disease; LUP = systemic lupus erythematosus; RA = rheumatoid arthritis; ST = ischemic stroke; LUNG = lung cancer; COPD = chronic obstructive pulmonary disease; T2D = Type 2 diabetes; LIV = Liver disease; IHD = ischemic heart disease, Breast = breast cancer; Colorectal = colorectal cancer; VD = vascular dementia).

Among the 225 protein-disease pairs, we replicated known causal relationships such as proprotein convertase subtilisin/kexin type 9 (PCSK9) and ischaemic heart disease^27^, and apolipoprotein E (APOE) and Alzheimer’s disease^28^. Other protein-disease pairs with significant causal associations in this study and prior evidence for causality included tumor necrosis factor receptor superfamily member 6B (TNFRSF6B) and inflammatory bowel disease (IBD)^29^, peptidylglycine alpha-amidating monooxygenase (PAM) and type 2 diabetes^30^, and transforming growth factor beta 1 (TGFB1) and ischaemic heart disease^31^. Sortilin1 (SORT1) had robust and significant evidence for causal effects on type 2 diabetes in this study and its potential causal role has not been previously identified.

We assessed the disease specificity of causal drivers identified in MR compared to the non-causal, disease-associated predictors. We found that hundreds of non-causal predictor proteins were associated with multiple diseases in the Cox PH models; several were associated with 9 or more diseases (e.g., amphiregulin (AREG), ribonuclease A family member k6 (RNASE6), and CUB domain-containing protein 1 (CDCP1)). The diseases associated with these highly non-specific proteins are involved in a broad range of mechanisms and pathways (**Figure 2b**). In contrast, most of the causal driver proteins (172/198, 87%) were associated with 1 disease. On average, this group of proteins was associated with 1.1 diseases, while the disease-associated proteins lacking MR support were associated with an average of 3.5 diseases.

### Many protein-disease associations are driven by smoking

Given the widespread sharing of non-causal predictor proteins across diverse diseases, we next tested the hypothesis that these proteins broadly associate with incident disease due to their associations with environmental risk factors, and thus reflect environmental impacts without directly causing disease. Previously developed proteomic scores for the incident disease outcomes showed better or similar prediction performance compared to models comprised of age, sex, and other lifestyle factors, including BMI, alcohol intake, social deprivation, educational attainment, physical activity, and smoking status^7^, pointing to a potential role of the plasma proteome in partially capturing the effects of these risk factors. For lung cancer and COPD specifically, we observed that incremental models with disease proteomic scores had minimal improvement beyond smoking status (**Extended Data** Fig. 2). Previous literature, including studies using small, targeted microarrays, suggests that smoking may alter plasma protein levels^32,33^. Thus, we further investigated the relationship between the plasma proteome and smoking, a well-known environmental risk factor with large effects on many common complex diseases^34^.

To first gauge the overall effects of smoking on the plasma proteome, we characterized the relationships between self-reported smoking status and the 2,923 proteins measured in the UKB-PPP cohort, adjusting for age and sex variables. 1,673 proteins (57%) were significantly associated with smoking status (Bonferroni-adjusted *P*-value < 1.7 x 10^-5^) (**Supplementary Table 5**). 30 proteins measured in the Olink panel mapped to 21 plasma proteins previously found to be associated with smoking^32^ (**Supplementary Table 6**), and all were significantly associated with smoking in these analyses.

We then evaluated the effects of smoking on the disease-associated proteins specifically. All 178 proteins associated with lung cancer in the Cox PH models were significantly associated with smoking, as were between 82-97% of proteins associated with all other diseases examined (**Figure 3a**), a notable enrichment beyond the proportion of all measured plasma proteins associated with smoking. Additionally, we found that all proteins associated with 8 or more diseases in the Cox PH models were smoking-associated, compared to smaller proportions of the more disease-specific proteins (**Figure 3b**).

**Figure 3.**
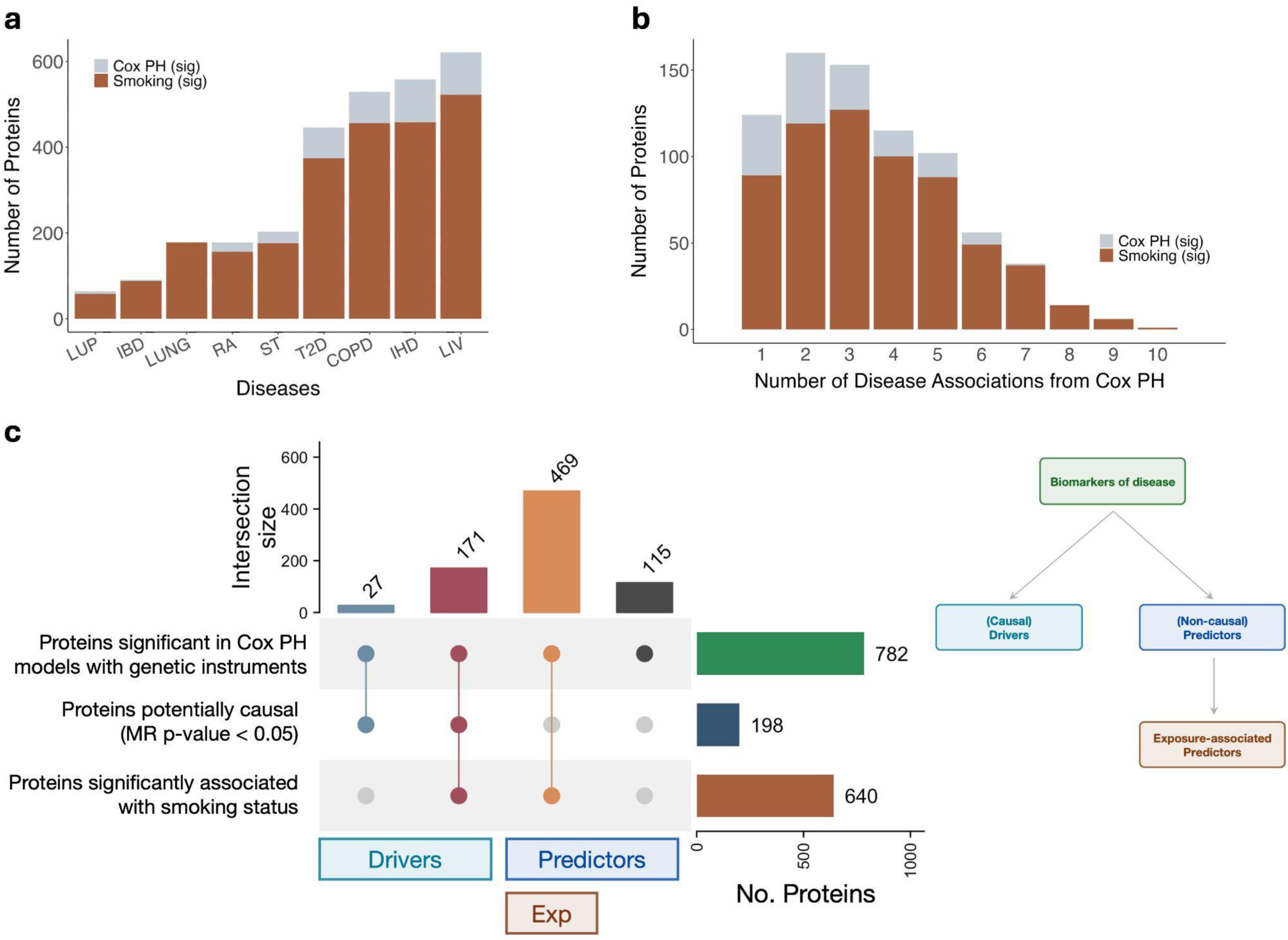
Smoking associations categorize many biomarkers as exposure-associated predictors. **a,** Counts of proteins significantly associated with diseases in Cox PH models, with counts of the subset of these proteins significantly associated with smoking status (across the entire UKB dataset) highlighted by orange bars. Diseases with more than 20 significant protein associations are shown (IBD = inflammatory bowel disease; LUP = systemic lupus erythematosus; RA = rheumatoid arthritis; ST = ischemic stroke; LUNG = lung cancer; COPD = chronic obstructive pulmonary disease; Diab = Type 2 diabetes; LIV = Liver disease; IHD = ischemic heart disease). **b,** Counts of proteins significantly associated with 1 to 10 diseases in the Cox PH models, with counts of the subset of proteins in each category significantly associated with smoking status highlighted by orange bars (Bonferroni-adjusted *P*-value < 1.7 x 10^-5^). **c,** UpSet plot showing the classification of 782 proteins significantly associated with a disease (Bonferroni-adjusted *P*-value < 0.05/(19 diseases x 2,923 proteins) = 9.00 x 10^-7^ from Cox PH associations) for which a genetic instrument existed. Significant association with smoking status was determined based on Bonferroni-adjusted *P*-value < 0.05/2,923 = 1.7 x 10^-5^. Groups delineated as causal drivers, non-causal predictors, and exposure-associated predictors are indicated by bars on the bottom, and illustrated in the schematic on the right.

After adjusting for self-reported smoking status in the Cox PH models, 8,326 out of 9,308 (89%) protein-disease pairs remained significant (**Supplementary Table 7**). Lung cancer showed the most substantial drop in the number of associated proteins after adjustment (from 324 to 33 proteins, representing a 90% decrease); COPD also showed a large decrease (from 1,501 to 1,250 proteins, representing a 17% decrease). Of note, most of the remaining lung cancer-associated proteins (28/33, 85%) had significantly attenuated hazard ratios after smoking adjustment (Wald test, p-value < 0.05), with a mean attenuation of 48.3% of the |*log*(*HR*)|. Taken together, these results indicate that although adjusting for smoking status yields large attenuations in the number of associated proteins for some diseases, basic measures of smoking may fail to capture the full effects of smoking in protein-disease associations.

Given the clear biological impacts of smoking, we then sought to systematically distinguish causal driver proteins from exposure-associated predictor proteins, focusing on the 782 disease-associated proteins with valid genetic instruments. We found that the vast majority of these proteins (N=640) were significantly associated with smoking status. Moreover, most of these smoking-associated proteins (N=469) showed no evidence for causal effects on disease from the *cis*-MR analyses (**Figure 3c**). Several proteins (N=171) were both associated with smoking and nominated by MR, indicating that these proteins may represent (or be correlated with) causal mediators between smoking and incident disease.

Another group of disease-associated proteins were neither associated with smoking nor showed evidence for causality (**Figure 3c**). The majority of proteins in this group (86/115, 75%) were associated with 1 to 3 diseases in the Cox PH models, potentially representing a group of proteins enriched for non-causal, disease-specific predictors tagging incident disease processes. For example, 5 proteins in this group were associated with only COPD, including a transmembrane activin inhibitor (BAMBI) that was previously found to be involved in Th17/Treg pathway imbalances in COPD patients^35^.

Based on these groupings, we characterized the proteins selected in our previously developed proteomic scores for COPD and lung cancer^7^. 47 proteins in the COPD score had at least one genetic instrument, and all except 2 (CLSTN2 and LYPD8) were significantly associated with smoking; 10 proteins additionally showed evidence for causal effects. In the lung cancer proteomic score, 5 of the 6 proteins had at least one genetic instrument, and all 5 were significantly associated with smoking; carcinoembryonic antigen (CEACAM5) additionally showed suggestive evidence for causality. These disease proteomic scores appear to mostly capture exposure-associated predictor proteins and not causal drivers, which aligns with our previous finding that they have diminished predictive utility when smoking status is included in the prediction models (**Extended Data** Fig. 2). Additionally, the proteomic score for lung cancer had limited prediction performance over the baseline model of age and sex in individuals that reported never smoking compared to previous and current smokers (ΔAUC between baseline plus lung cancer ProteinScore and baseline only = 0.02, 0.06, and 0.05 in never, previous, and current smokers, respectively).

### Proteomic score quantifies smoking behavior and history

The widespread effects of smoking on the plasma proteome indicate that plasma proteins themselves may serve as precise, quantitative measures of the cumulative biological effects of smoking and other exposures. To assess the utility of the plasma proteome as a quantitative readout of smoking, we developed a smoking protein score (SmokingPS). Since many of the measured proteins are likely correlated, we performed variable selection via LASSO on the protein levels of current and never smokers in the UKB (N=14,585) (**Extended Data** Fig. 3), identifying 550 proteins that we combined into the SmokingPS. This score predicted smoking status with high accuracy (AUC = 0.955 [95% CI = 0.949-0.961]) in a hold-out set of UKB participants (N=14,586). We replicated the score in another biobank, FinnGen^36^, with plasma protein measurements from the Olink Explore panel in 1,990 participants, 1,863 of whom had smoking status information. The SmokingPS demonstrated good but lower ability to discriminate current vs. never smoking status in FinnGen participants (AUC = 0.844 [95% CI = 0.814-0.875]), and the SmokingPS distributions showed some separation between current/previous smokers and never smokers (**Extended Data** Fig. 4a). We note that because smoking status information was collected at recruitment several years before proteomic sampling in FinnGen, it was not possible to distinguish between current and previous smokers for this analysis.

We compared the distributions of the SmokingPS across individuals in UKB-PPP who reported current, previous, and never smoking status. As expected, the previous smokers had a SmokingPS distribution between the current and never smokers (**Figure 4a**). We then stratified the previous smokers by years since smoking cessation and pack years, which captures both quantity and frequency of smoking. As years since smoking cessation increased, the distributions of the SmokingPS in previous smokers shifted towards the never smokers (**Figure 4b**). As illustrated by the SmokingPS distributions across individuals who stopped smoking recently, the smoking-associated proteome starts to revert back to non-smoking levels within a couple years of smoking cessation; this pattern is also observed from the levels of the top-weighted proteins in the SmokingPS, averaged across bins of years since smoking cessation in former smokers (**Extended Data** Fig. 5). Similarly, those who smoked fewer pack years had SmokingPS distributions closer to the never smokers (**Extended Data** Fig. 4b). Stratifying current smokers by pack years reflected the same trend – individuals who reported fewer pack years had SmokingPS distributions closer to, or overlapping, that of previous smokers (**Figure 4c**). The average number of cigarettes smoked per day among current smokers ranged from 8.6 in the lowest quantile of SmokingPS distribution to 20.1 in the highest quantile, underlining that the amount of smoking has a dose-response impact on plasma protein levels (**Extended Data** Fig. 4c).

**Figure 4.**
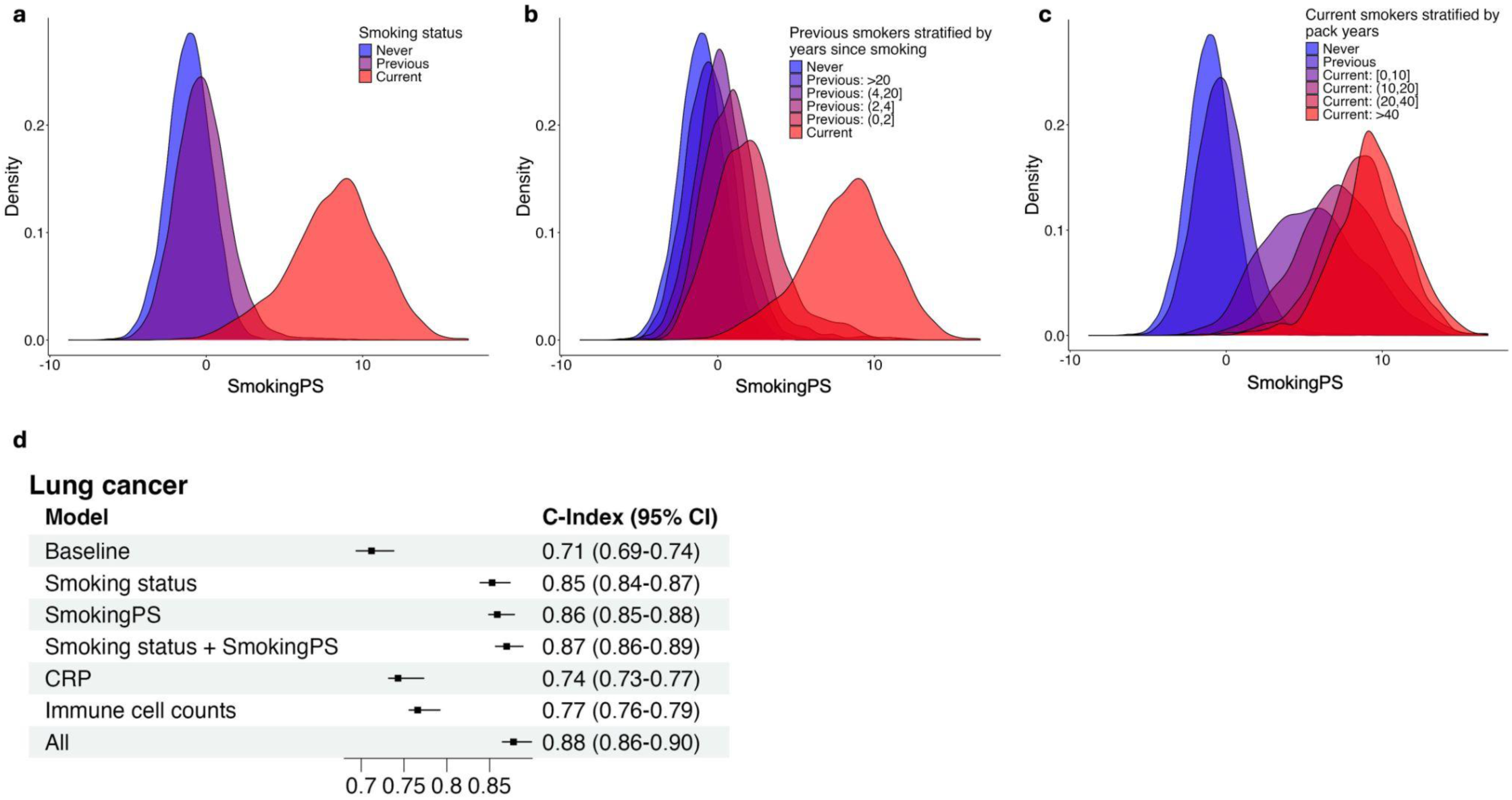
SmokingPS captures different smoking measures and predicts incident disease. **a,** Density plot of SmokingPS with individuals stratified by self-reported smoking status. **b,** Density plot of SmokingPS with self- reported previous smokers stratified by number of years since smoking cessation. **c,** Density plot of SmokingPS with self-reported current smokers stratified by pack years, calculated as number of cigarettes smoked per day, divided by twenty, multiplied by number of years smoking. **d,** Associations between incident lung cancer (cases = 405, controls = 46,968) and models with various predictors, shown on *y*-axis, using Cox PH. Baseline model includes age, sex, age^2^, age × sex, age^2^ × sex, and first 10 genetic PCs. All models following baseline include these variables, as well as the predictor listed. “CRP” indicates C-reactive protein. “Immune cell counts” include neutrophil, eosinophil, basophil, monocyte, lymphocyte, and white blood cell counts. “All” indicates the baseline variables, smoking status, SmokingPS, CRP, and immune cell counts. C-index is shown on the *x-*axis and listed alongside 95% confidence intervals.

We further tested the value of this SmokingPS for disease prediction (**Figure 4d**, **Extended Data** Fig. 6). Among the 19 incident disease outcomes, adding smoking status to baseline variables of age, sex, and genetic principal components (PCs) resulted in the greatest improvement in performance for lung cancer, COPD, and liver disease (delta C-index between baseline model vs. baseline with smoking status: 0.141, 0.083, and 0.024, respectively). Adding the SmokingPS to the baseline model with smoking status further improved performance for these three diseases (delta C-index between baseline with smoking status vs. with smoking status and SmokingPS: 0.017, 0.011, and 0.008 for lung cancer, COPD, and liver disease), indicating that the plasma proteome likely provides a more quantitative biological readout of smoking exposure than self-reported smoking status for some diseases.

Of the individuals with incident lung cancer diagnoses (N = 412), 58 were self-reported never smokers. The SmokingPS showed better prediction of lung cancer incidence over the baseline model in current smokers (delta C-index = 0.044, p-value = 0.069) compared to in the never smokers (delta C-index = 0.013, p-value = 0.71) (**Extended Data** Fig. 7c). Additionally, distributions of the SmokingPS between never smokers with and without lung cancer were not significantly different (two-sample Kolmogorov-Smirnov test *D* = 0.15, p-value = 0.11) (**Extended Data** Fig. 7b). We then considered the full set of non-smokers, and leveraged the breadth of exposure data collected in the UKB to investigate other factors that the SmokingPS may be associated with in never smokers. Across 71 environmental and lifestyle factors covering a range of variables (e.g. secondhand smoke exposure, physical and social activities, air pollution measures) (**Supplementary Table 8**), the SmokingPS was significantly associated with a few smoking-related factors, such as “attends pub/club”, “exposure to tobacco smoke at home”, and “number of smokers in household” (**Supplementary Table 9**).

In additional sensitivity analyses, we compared the SmokingPS to commonly-used clinical biomarkers, C-reactive protein (CRP) and immune cell counts (**Figure 4d**, **Extended Data** Fig. 6a). For lung cancer and COPD, the SmokingPS outperformed both CRP and immune cell counts (delta C-index between baseline with SmokingPS vs. with CRP: 0.116 and 0.051 for lung cancer and COPD, respectively; delta C-index between baseline with SmokingPS vs. with immune cell counts: 0.093 and 0.015). Combining CRP and immune cell counts with both smoking status and the SmokingPS resulted in the best performing models for lung cancer, COPD, and liver disease, indicating that each measure may capture unique aspects of disease risk.

### Generalization of paradigm: proteomic score for alcohol use

Having demonstrated that the proteome can effectively serve as a quantitative measure of disease-relevant environmental risk factors using smoking as an example, we sought to explore whether this approach was generalizable to other environmental exposures. We tested associations between the plasma proteome and alcohol intake status, and found that 1,228 (42%) of proteins were significantly associated with alcohol use (Bonferroni-adjusted P-value < 1.7 x 10-5) (**Supplementary Table 10**). We developed a proteomic score for alcohol intake (AlcoholPS), trained on the protein levels of UKB-PPP individuals who reported never drinking and daily drinking. The score consisted of 474 proteins selected by LASSO, and predicted current vs. never drinking status in a hold-out set of UKB participants with high accuracy (AUC = 0.975). Although 134 (28%) proteins overlapped with those in the SmokingPS, the AlcoholPS distributions did not stratify according to smoking status and vice versa (two-sample Kolmogorov-Smirnov test for AlcoholPS: *D =* 0.16, p-value < 2.2e- 16 for never vs. previous smokers; *D =* 0.14, p-value < 2.2e-16 for never vs. current smokers; *D =* 0.05, p-value = 3.96e-09 for previous vs. current smokers; and SmokingPS: *D =* 0.17, p-value < 2.2e-16 for never vs. previous drinkers; *D =* 0.17, p-value < 2.2e-16 for never vs. current drinkers; *D =* 0.07, p-value = 1.73e-08 for previous vs. current drinkers) (**Extended Data** Fig. 8a-b), indicating that the two scores are largely independent.

Distributions of the AlcoholPS in self-reported current drinkers stratified by number of drinks per week and month (**Fig. 5a**). Additionally, we stratified current drinkers by the number of grams of alcohol intake per day, derived from several alcohol intake variables in the UKB (**Methods**), and similarly observed gradual shifts in the protein score distribution as the alcohol intake quantities increased (**Fig. 5b**). Given differences in the ratio of drinkers vs. never-drinkers in males (653 never-drinkers; 5,233 current drinkers) and females (1,498 never-drinkers; 3,891 current drinkers), we performed additional sex-stratified sensitivity analyses. AlcoholPSs trained separately in males and females similarly stratified according to alcohol intake frequency and amount (**Extended Data** Fig. 8c-f), indicating that the AlcoholPS is not merely detecting sex-specific proteins. The AlcoholPS derived from matching sample sizes of male and female never and daily drinkers in the training cohort (**Methods**) had similar performance in the hold-out set (AUC = 0.969), and 293/408 (72%) of the proteins in this AlcoholPS overlapped with the AlcoholPS trained on all individuals without downsampling.

**Figure 5.**
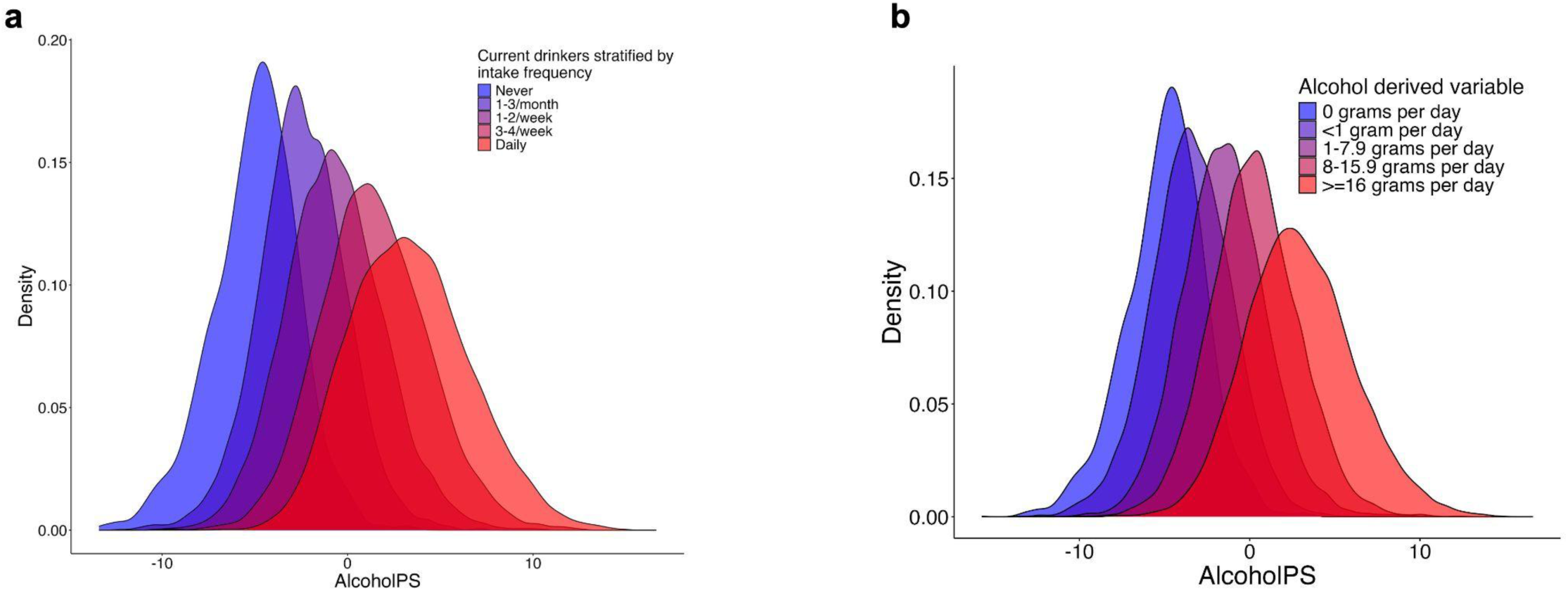
AlcoholPS captures frequency and amount of alcohol intake in current drinkers. **a,** Density plot of AlcoholPS in current drinkers stratified by self-reported number of drinks per week or month. Self-reported never drinkers shown for reference. **b,** Density plot of AlcoholPS in current drinkers stratified by derived grams of alcohol intake per day.

We additionally evaluated the ability of the AlcoholPS to predict clinical biomarkers commonly used to assess liver function, including alanine aminotransferase (ALT), aspartate aminotransferase (AST), gamma-glutamyl transferase (GGT), and bilirubin. The AlcoholPS outperformed self-reported alcohol intake status in daily and never drinkers for AST (incremental *R^2^*: 0.005 vs. 0.0009), GGT (0.027 vs. 0.004), and bilirubin (0.02 vs. 0.01), as well in current and never drinkers (**Extended Data** Fig. 9).

## Discussion

Our study shows that a substantial proportion of plasma protein-disease associations identified in observational studies are the result of systemic effects of environmental risk factors on protein expression and do not reflect causal relationships between those proteins and disease. Instead, our results suggest that much of the predictive value of the disease-associated proteome stems from its association with environmental impacts, which have been historically challenging to evaluate consistently across studies using objective and quantitative approaches. This has significant implications for the clinical translation of disease risk estimates and biomarkers derived from plasma proteomics, as it indicates that in many cases the proteins may be capturing the effects of an upstream exposure driving changes in protein expression rather than directly contributing to disease onset. Our approach can be applied to numerous diseases and exposure types, helping delineate how plasma proteomic information can inform health interventions.

Associations between protein levels and disease incidence observed in epidemiological studies are susceptible to confounding and other biases. We leveraged MR to determine which subset of protein-disease associations may represent direct, causal effects of proteins on disease onset. Many prior studies have focused on identifying causal proteins for specific diseases^10,13,14^, whereas our study investigated 19 different diseases that span common metabolic diseases (e.g. type 2 diabetes), immune-mediated diseases (e.g. rheumatoid arthritis and IBD), and cancers to discern potential patterns or differences in the proportions of causal proteins identified between traits. Across diseases, we found only 8% of protein-disease pairs tested had any suggestive evidence for a causal relationship. The small proportion of putatively causal protein drivers identified in this study is consistent with findings from disease-specific studies, as well as two recent phenome-wide MR studies: one study found that less than 1% of the tested protein-phenotype pairs across 355 phenotypes were putatively causal^20^, and another study, which included different diseases and filters, similarly found less than 1% of significant protein-disease pairs were putatively causal using *cis*-pQTLs^9^. Despite the low numbers of MR-nominated proteins, we confirm that this group of proteins includes more disease-specific signals, rather than broadly disease-associated proteins. Though MR-nominated proteins represent a small fraction of the proteome, their potential causal roles are critical to mechanistically characterize as they may constitute promising therapeutic targets.

In the Cox PH models associating proteins with incident disease, we observed that many proteins were associated with multiple, often unrelated, diseases. One explanation is that environmental factors that impact shared pathways underlying many diseases also have widespread effects on the plasma proteome. In this study we explored this scenario and focused on smoking as an example of a known environmental risk factor for many diseases with persistent biological effects^37^. By integrating the MR and smoking analyses, we determined that for many proteins tested for causal effects on disease, their associations with incident disease outcomes are merely coincidental due to their independent associations with smoking. Our partitioning framework illustrates a strategy for honing in on the proteins that may be most relevant to disease – whether as putative therapeutic targets for pharmacological intervention (i.e. causal drivers), as quantitative sensors of environmental influences on disease (i.e. exposure-associated predictors), or alternatively as biomarkers of disease-specific processes beginning before diagnosis (i.e. non-causal predictors without smoking or other environmental exposure association). We note that our study is not an exhaustive investigation of potential environmental perturbations, and thus future work applying our approach to other environmental risk factors will enable further refinement of these groups. Based on our finding that disease-based proteomic scores for lung cancer and COPD were largely composed of smoking-associated predictors, other disease-based proteomic scores may similarly include non-specific signals of other exposures that have extensive effects on the plasma proteome.

We developed a proteomic score for smoking that demonstrates the potential of the plasma proteome to provide precise biomarkers of environmental risk factors. Distributions of the SmokingPS among previous and current smokers demonstrated the dynamic nature of the plasma proteome, sensitive to not only smoking status but also intensity and duration of smoking. This points to the ability of the plasma proteome to serve as an accurate and detailed proxy for smoking behavior and history in addition to, or even instead of, questionnaire data. For example, we identified a group of individuals in UKB who were labeled as current smokers but had a SmokingPS distribution overlapping that of previous smokers; other questionnaire data in the UKB about smoking cessation revealed that they were most likely previous smokers. Potentially of even greater utility is the ability of the SmokingPS to capture and quantify heterogeneity among current smokers in terms of number of cigarettes smoked or total years of smoking, particularly for datasets that do not have questionnaire data as detailed as in the UKB.

The ability of the SmokingPS to discriminate between current and never smokers (AUC=0.96) was similar although slightly lower compared to blood-based DNA methylation predictors of smoking, which were able to discriminate between current and never smokers with an AUC of 0.98^38^. Studies have found that methylation levels of most cytosine–phosphate–guanine sites (CpGs) revert to that of never smokers within 5 years of smoking cessation^39,40^, and the SmokingPS of previous smokers similarly resembled that of never smokers after around 4-5 years of smoking cessation. However, some CpGs have been found to not revert to the levels of never smokers even after 30 years of smoking cessation. In this study, we observed that proteins with the largest weights in the SmokingPS returned back to never-smoker levels within 5 years. Given the complexity and slower pace of epigenetic reprogramming^41^ and the greater biological interpretability of protein levels, plasma proteins may offer a more sensitive and mechanistically informative indicator of smoking. Nevertheless, large-scale datasets with both types of measurements will be needed to better understand the dynamics between epigenetic and proteomic biomarkers.

The SmokingPS was also associated with incidence of several diseases, particularly diseases for which smoking is a known risk factor. The addition of the score to the smoking status variable offered modest improvements in prediction over the smoking status variable alone. This underscores that for diseases like lung cancer and COPD, the predictive ability of the plasma proteome likely lies in its ability to accurately quantify smoking effects. Sample sizes were too small in the UKB-PPP cohort to compare disease-based proteomic scores trained in non-smoking lung cancer and COPD patients, but this approach may reveal insights into the biology of these diseases unrelated to smoking, or the effects of other non-smoking exposures such as respiratory infections^42^ and poor cooking ventilation^43^.

The AlcoholPS demonstrated similar effectiveness in quantifying alcohol intake behaviors, suggesting that the value of the proteome in this context is not limited to smoking. Self-reported alcohol intake has been shown to be particularly subject to misreporting and confounding, which has resulted in biased assessments of the effects of alcohol consumption in observational studies^44^. Thus, there is a critical need for more objective proxies of alcohol intake, and our study provides evidence that the plasma proteome may fill this gap. However, we note that we did not observe associations between alcohol status and incident diseases in this study, and thus we did not pursue disease prediction analyses with the AlcoholPS. Alcohol-related disorders, such as alcoholic pancreatitis (N = 94), alcoholic cardiomyopathy (N = 8), cirrhosis (N = 43), and oropharyngeal cancers (N = 9), had limited sample sizes of incident outcomes in the UKB-PPP cohort, precluding prediction analyses using the AlcoholPS.

We note several limitations. First, although we obtained the largest GWAS publicly available without UKB data for estimating the effects of genetic instruments on disease outcomes, limits in sample sizes may have limited statistical power for estimating effects in MR. Second, we adopted several strategies to ensure the robustness of the MR results, but violations of model assumptions are still possible; thus, potential causal drivers identified in this study will require further validation. Third, we utilized data from only European ancestry populations for the MR analyses, potentially limiting generalizability of the findings to other groups, although causal genetic effects are largely shared across populations^45,46^. Fourth, the proteins captured by these proteomic assays are not a completely random subset of the proteome; therefore, proteins involved in certain disease-specific pathways may not be represented or below limits of detection, and proteins involved in general disease activity may be disproportionately represented. Finally, we did not explore the possibility that some of the disease-associated proteins may be biomarkers of early disease processes that begin prior to diagnosis due to limitations in EHR data. A key challenge for deeply curated clinical cohorts will be distinguishing between effects from unmeasured environmental factors and those arising from uncharacterized disease mechanisms.

In conclusion, we highlight the environment as a prominent component of the reported predictive power of the disease-associated plasma proteome for patient outcomes. This underscores the challenge of interpreting proteomic data, given that environmental exposures, alongside inherited genetic variation and ongoing disease processes, have substantial impacts on plasma proteins. Drawing insights into disease-specific mechanisms from this data will require systematic characterizations to separate true causal disease signals from more general reflections of smoking or other factors. This is critical for not only understanding disease biology but also improving disease prediction, since models with more disease-specific proteomic biomarkers may be more portable across populations. Especially as the breadth of protein measurements are increasing at decreasing cost, clarifying the roles of plasma proteomic measurements from these biobank datasets will be an important step towards clinical translation.

Our work also suggests that proteomic assays may open up a path toward measuring the impacts of the environment on human health and disease. Assessing genetic risk is now largely straightforward, thanks to assays that enable consistent, reproducible measurements across studies. However, this has not been the case for studies of environmental risk factors, where data tends to be collected using non-standardized questionnaires and a diversity of other methods. Our findings point to the potential role of proteomic assays as a way to extend insights from smaller-scale studies on environmental exposures to other studies lacking comparable data, as well as develop quantitative biomarkers for exposures that may not have existing biological readouts. While our study focused on evaluating the aggregate predictive power of plasma proteins for disease-related exposures, complementary studies have focused on partitioning the variance in individual protein levels explained by exposures vs. genetics^24,25^. Although these studies show that only a small proportion of proteins are more influenced by non-genetic factors than genetics, we illustrated that in aggregate plasma proteins are highly accurate readouts of lifestyle factors (AUC = 0.96 for smoking, 0.98 for alcohol intake) because such a large proportion of the proteins are perturbed by these factors. Well-powered proteomic cohorts with detailed environmental measures and longitudinal health records are needed to comprehensively disentangle the effects of the environment vs. other factors like early disease processes on the disease-associated plasma proteome.

## Methods

### Proteomic profiling in the UK Biobank

The UK Biobank Pharma Proteomics Project (UKB-PPP) is a precompetitive biopharmaceutical consortium formed with the goal of collecting and characterizing the plasma proteomic profiles of participants from the UKB^47^, a population-based cohort comprising approximately 500,000 individuals from the United Kingdom. The UKB has been described in Bycroft et al.^47^ and details on the data available in the UKB can be found at https://biobank.ndph.ox.ac.uk/showcase/. 54,219 participants were selected for the UKB-PPP cohort; we restricted the sample used for these analyses to participants within the cohort who were randomly sampled from the main UKB population. The Olink Explore 3072, an antibody-based proximity extension assay, was used to measure the abundance of protein analytes in each plasma sample. Measurements were provided in the Normalized Protein eXpression (NPX) values on a log_2_ scale. Full details on sample selection, the Olink assay, and data processing and quality control are described in Sun et al.^6^. We excluded three proteins missing in >10% of the sample (CTSS, NPM1, and PCOLCE), and imputed protein expression values of the remaining proteins using the miceforest package in Python. All proteins except those missing in >30% of participants were used as predictors for the imputation of each protein. We imputed a single dataset using a maximum of five iterations. All other parameters were left at default values. After imputation, proteomic data were normalized separately within each cohort by first rescaling values to be between 0 and 1 using MinMaxScaler() from scikit-learn and then centering on the median. The final dataset consisted of 2,923 proteins measured in 45,438 individuals.

### Disease outcomes and other phenotypes in the UK Biobank

Codes and data used to define prevalent and incident disease in the UKB are detailed in Supplementary Table 20 from our previous publication^48^. Diagnoses and date of first diagnosis for all diseases in the UKB were ascertained using ICD diagnosis codes and corresponding dates of diagnosis taken from linked hospital inpatient, primary care and death register data. If a participant did not have a diagnosis code in hospital inpatient or primary care records, but the code was listed as a primary or secondary cause of death, then they were coded as a case with the date of diagnosis as the date of death. Primary care read codes were converted to corresponding ICD diagnosis codes using the lookup table provided by the UKB. Linked hospital inpatient, primary care and cancer register data were accessed from the UKB data portal on 22 February 2024, with a censoring date of 31 October 2022; 31 August 2022 or 31 May 2022 for participants recruited in England, Scotland or Wales, respectively (8–16 years of follow-up). Detailed information about the linkage procedure national registries for mortality and cause of death information in the UKB is available online (https://biobank.ctsu.ox.ac.uk/crystal/refer.cgi?id=115559) with. Mortality data were accessed from the UKB data portal on 22 February 2024, with a censoring date of 30 November 2022 for all participants (12–16 years of follow-up).

We investigated the 23 diseases studied in Gadd et al.^7^, which represent a selection of leading age-related diseases: liver disease, systemic lupus erythematosus, type 2 diabetes, amyotrophic lateral sclerosis, Alzheimer’s dementia, endometriosis, COPD, inflammatory bowel disease, rheumatoid arthritis, ischemic stroke, Parkinson’s disease, vascular dementia, ischemic heart disease, major depressive disorder, schizophrenia, multiple sclerosis, cystitis, and lung, prostate, breast, gynecological, brain/central nervous system and colorectal cancers. Gynecological and breast cancer, endometriosis and cystitis were female-stratified; prostate cancer was male-stratified. For these diseases, sex was not included as a covariate in the Cox PH models.

For smoking associations, self-reported smoking status (field ID 20116) was coded as a categorical dummy variable (0 = never, 1 = previous, 2 = current). Individuals who were labeled as never smokers in this field but reported being current or previous tobacco smokers (field ID 22506) were excluded from all analyses. Additional smoking variables used included: pack years (field ID 20161), cigarettes smoked per day in current smokers (field ID 3456), cigarettes smoked per day in previous smokers (field ID 2887), age stopped smoking from questionnaire data (field ID 2897), and age stopped smoking from medical records (field ID 22507). Number of years since smoking was computed based on the baseline age and age stopped smoking variables as follows: first, individuals with an age reported in either of the age stopped smoking variables were included; if individuals had ages reported in both fields which differed by 6 or fewer years, they were included and the average age across the two fields was used; finally, years since smoking was calculated by subtracting the age of smoking cessation from the baseline age.

For sensitivity analyses, C-reactive protein (CRP) and immune cell counts were also extracted from UKB (CRP: field ID 30710; basophil counts: field ID 30160; eosinophil counts: field ID 30150; lymphocyte counts: field ID 30120; monocyte counts: field ID 30130; neutrophil counts: field ID 30140; white blood cell counts: field ID 30000). CRP levels were natural log transformed; the dataset did not have outliers, defined as 4 standard deviations from the mean. White blood cell counts were transformed to Z-scores and outliers 4 standard deviations from the mean were excluded (N outliers = 76). Rank-based inverse normal transformation was performed on all other immune cell count phenotypes.

In never smokers, we additionally collated a selection of 71 exposures, covering behavioral (e.g. exercise habits, media use), lifestyle (e.g. social activities, transportation) and environmental (e.g. pollution measures, traffic density) domains. The selection was partially curated based on Choi et al.^49^. The full list of variables can be found in **Supplementary Table 8.**

For alcohol analyses, self-reported alcohol status (field ID 20117) was coded as a categorical dummy variable (0 = never, 1 = previous, 2 = current), and self-reported alcohol intake (field ID 1558) was also coded as a categorical dummy variable (4 = 1-3 drinks/month, 3 = 1-2 drinks/week, 2 = 3-4 drinks/week, and 1 = daily). Grams per day of alcohol intake were calculated using a method from a previously published paper in the UKB^50^. Liver biomarkers were extracted from UKB (ALT: field ID 30620; AST: field ID 30650; GGT: field ID 30730; Bilirubin: field ID 30840). Outliers, defined as 4 standard deviations from the mean, were excluded (ALT N outliers = 322; AST N outliers = 269; GGT N outliers = 425; bilirubin N outliers = 396). Rank-based inverse normal transformation was performed on all liver biomarkers.

### Cox PH models

To identify which proteins were significantly associated with incident disease, we ran Cox proportional hazards (PH) models between each protein and incident disease outcome using the survival package (v3.4-0) in R^51^. Protein levels were inverse rank normalized for these models. The models were adjusted for age at baseline and sex; for sex-stratified outcomes, only age was included as a covariate. To assess significance, we used a Bonferroni-adjusted *P*-value threshold for multiple testing based on the 23 disease outcomes and 2,923 proteins tested (*P*-value < 0.05/(23 x 2923) = 7.41 x 10^-7^).

### Mendelian Randomization

To determine the subset of protein-disease associations identified by the Cox PH models that also potentially represent causal relationships, we performed two-sample Mendelian Randomization (MR). We used protein quantitative trait loci (pQTLs) as genetic instruments, which had been previously identified from GWAS of the 2,941 protein analytes measured in 34,557 individuals of European ancestry in the UKB-PPP cohort^6^. We first mapped pQTLs to the GRCh38/hg38 build and selected any genome-wide significant pQTL (P-value < 5x10^-8^). We then excluded pQTLs located within the human major histocompatibility complex (MHC) region on chromosome 6 (positions 28510120 to 33480577), due to the complex LD structure in this region, and on the sex chromosomes. We additionally excluded pQTLs with MAF <= 0.005 and an INFO score <= 0.8.

We restricted our analyses to the *cis-*pQTLs, defined as those on the same chromosome and within 1Mb of the transcriptional start site of the protein-coding gene. SNPs were allowed to be *cis-*pQTLs for one or more proteins. To identify independent *cis-*pQTLs, we applied clumping in PLINK (v1.9b7-x86_64) for each protein, using an LD threshold of r2 <= 0.001 and a reference panel including the 34,557 individuals of European ancestry from the UKB-PPP GWAS of protein levels^6^. To prevent underflow, -log10(P-values) across proteins were scaled to the [0,1] range before clumping.

Genetic association data for the outcomes were selected based on exclusion of UKB data, to avoid overlap with the UKB-PPP cohort. 19 of the 23 disease outcomes had publicly-available GWAS that did not include UKB data and included only individuals of European ancestry, or were available in the FinnGen study^36^ (freeze 10). To boost power, we performed a meta-analysis of GWAS from FinnGen and Okada et al.^52^ for rheumatoid arthritis (RA) using the METAL software^53^. For outcome GWAS without minor allele frequency (MAF) information, which is required to perform Steiger filtering, we extracted MAF data from the non-Finnish European ancestry group (NFE) in gnomAD v4.1.0^54^. For the rheumatoid arthritis meta-analysis, the MAF used was computed as follows: (MAF_FinnGen_*N_FinnGen_ + MAF_gnomAD_NFE_*N_Okada_2014_)/(N_FinnGen_ + N_Okada_2014_), where FinnGen refers to the FinnGen GWAS for RA and Okada_2014 refers to Okada et al.^52^. The full list of outcome GWAS can be found in **Supplementary Table 2**.

For each outcome GWAS, we then harmonized the *cis-*pQTLs to ensure the effect allele of the SNPs in the exposure and outcome GWAS matched. We used the R package TwoSampleMR^55^ to perform the harmonization. We additionally excluded *cis-*pQTLs with *F-*statistics <= 10, and performed Steiger filtering to exclude variants with larger correlations with the outcome than the exposure. Ultimately, we tested 2,373 protein-disease pairs with genetic instruments that were significant (Bonferroni-adjusted *P-*value < (19*2923) = 8.97 x 10^-7^) in the Cox PH associations (**Supplementary Table 3**).

The Wald ratio test was applied to proteins with only one *cis-*pQTL, and the inverse-variance-weighted (IVW) method was applied to proteins with 2 or more *cis*-pQTLs. Additionally, for protein-disease pairs with more than 2 *cis*-pQTLs, MR-Egger was applied to test for horizontal pleiotropy. These tests were run using the MendelianRandomization package in R^56^. Heterogeneity statistics were also extracted from the IVW tests from the MendelianRandomization package. Scripts to perform genetic instrument harmonization and run the MR tests were adapted from https://github.com/globalbiobankmeta/multi-ancestry-pwmr^21^.

### Smoking associations

Associations between inverse rank normalized protein levels and self-reported smoking status in the UKB were tested using linear regression using the speedglm package^57^. All models were adjusted for age, sex, age x sex, age^2^, and age^2^ x sex. A Bonferroni-adjusted *P*-value threshold for multiple testing was used (*P*-value < 0.05/2923 = 1.7 x 10^-5^) to assess significance. The same approach was applied to test associations between the inverse rank normalized protein levels and self-reported alcohol status in the UKB.

To partition proteins based on the MR analyses and smoking associations, we first extracted all protein-disease pairs tested in the Cox PH models that were also tested in MR (i.e. had valid genetic instruments). We then took the subset of protein-disease pairs that reached significance in the Cox PH models based on a Bonferroni-adjusted *P-*value threshold (*P*-value < 0.05/(19 x 2923) = 8.97 x 10^-7^). The unique proteins across these protein-disease pairs (N = 782) were then carried forward and combined with the smoking association results to determine which of these proteins were significantly associated with smoking. Results were visualized and plotted in an UpSet plot using the ComplexHeatmap package (v2.15.4)^58^ in R. All above analyses were conducted using R v.4.4.0.

### Proteomic Score for smoking and alcohol intake

We developed a proteomic score for smoking using LASSO logistic regression in the R package glmnet (v4.1.8)^59^. We trained this score using the protein levels of UKB-PPP individuals who reported current and never smoking. We first randomly sampled 50% of this cohort to use for training and performed 10-fold cross-validation to select protein features and derive their weighting coefficients. We then generated scores in the remaining 50% of the cohort not used for training by computing the weighted sum of the levels of proteins selected in training. These scores were evaluated using AUC statistics calculated via the R package pROC (v1.18.5)^60^. We also evaluated the score, using the LASSO weights of the selected proteins, in participants from the FinnGen study with plasma protein measurements from Olink Explore and smoking status information (N current/previous smokers = 850 and N never smokers = 1,013).

To further evaluate the SmokingPS, we stratified the UKB-PPP cohort by years since smoking cessation in previous smokers, pack years in current and previous smokers, and cigarettes smoked per day in current smokers. Current smokers who reported an age stopped smoking or did not report cigarettes smoked per day were excluded from these analyses. Additionally, outliers with pack years or cigarettes smoked per day 4 standard deviations away from the mean were excluded.

We tested associations between the SmokingPS and incident disease outcomes (the 19 diseases that were tested in MR) using Cox PH models using the survival package (v3.4-0) in R^51^. For comparison, seven models were tested: (1) a baseline model including age, sex, age^2^, age × sex, age^2^ × sex, and first 10 genetic PCs, sourced from the Pan-UKB project^61^; (2) all covariates in the baseline model and smoking status; (3) all covariates in the baseline model and the SmokingPS; (4) all covariates in the baseline model, smoking status, and the SmokingPS; (5) all covariates in the baseline model and CRP; (6) all covariates in the baseline model and immune cell counts; (7) all covariates in the baseline model, smoking status, the SmokingPS, CRP, and immune cell counts. Results were plotted using the R package forestploter^62^. Associations between the SmokingPS and 71 environmental exposures were tested in never smokers using the glm function in R.

For the proteomic score for alcohol intake, we followed the same protocol used to develop and evaluate the SmokingPS. We trained this score using the protein levels of UKB-PPP individuals who reported never (N = 2,165) and daily (N = 9,185) drinking. We also developed sex-specific AlcoholPS, by first splitting individuals by sex and then training the scores separately in males (N daily = 5,233; N never = 653) and females (N daily = 3,891; N never = 1,498). We additionally matched the sample sizes of never and daily drinkers to that of the smaller sex-specific group (i.e. downsampled daily drinkers in males to the sample size of daily drinkers in females and never drinkers in females to the sample size of never drinkers in males), and evaluated the score trained on the combined dataset of males and females with matching sample sizes. To evaluate the AlcoholPS, we stratified the current drinkers by alcohol intake frequency, as well as bins of grams of alcohol intake as defined by the derived alcohol intake variable.

## Data availability

Individual-level data from the UK Biobank can be accessed via application at https://www.ukbiobank.ac.uk/. We accessed UK Biobank data under application 31063. GWAS summary statistics can be accessed as described in their respective papers in Supplementary Table 2 and in the FinnGen study^36^. Weights for the proteomic scores will be made available upon publication. Further information and requests for resources should be directed to and will be fulfilled by the lead contact, Kristin Tsuo, upon reasonable request. Code used for data preparation and analysis will be made available on GitHub before journal publication.

## Author Contributions

Study design, K.T., A.R.M., and M.J.D.; data analysis, K.T., M.A.A., D.G., D.B.; interpretation of results, K.T., A.R.M., M.J.D., M.A.A., M.K., Z.Z., R.E.M., C.F., H.H., B.S., and C.C.; writing, K.T., A.R.M., and M.J.D.

## Data Availability

Individual-level data from the UK Biobank can be accessed via application at https://www.ukbiobank.ac.uk/. We accessed UK Biobank data under application 31063. GWAS summary statistics can be accessed as described in their respective papers in Supplementary Table 2 and in the FinnGen study. Weights for the proteomic scores will be made available upon publication. Further information and requests for resources should be directed to and will be fulfilled by the lead contact, Kristin Tsuo, upon reasonable request.

## Acknowledgements

A.R.M is funded by NIH U01HG011719 as well as Broad Institute Next Gen and Merkin awards. K.T. is funded by F31HL167378 and supported by the ECOR Claflin Award to A.R.M.

---

**Extended Data Fig. 1.**
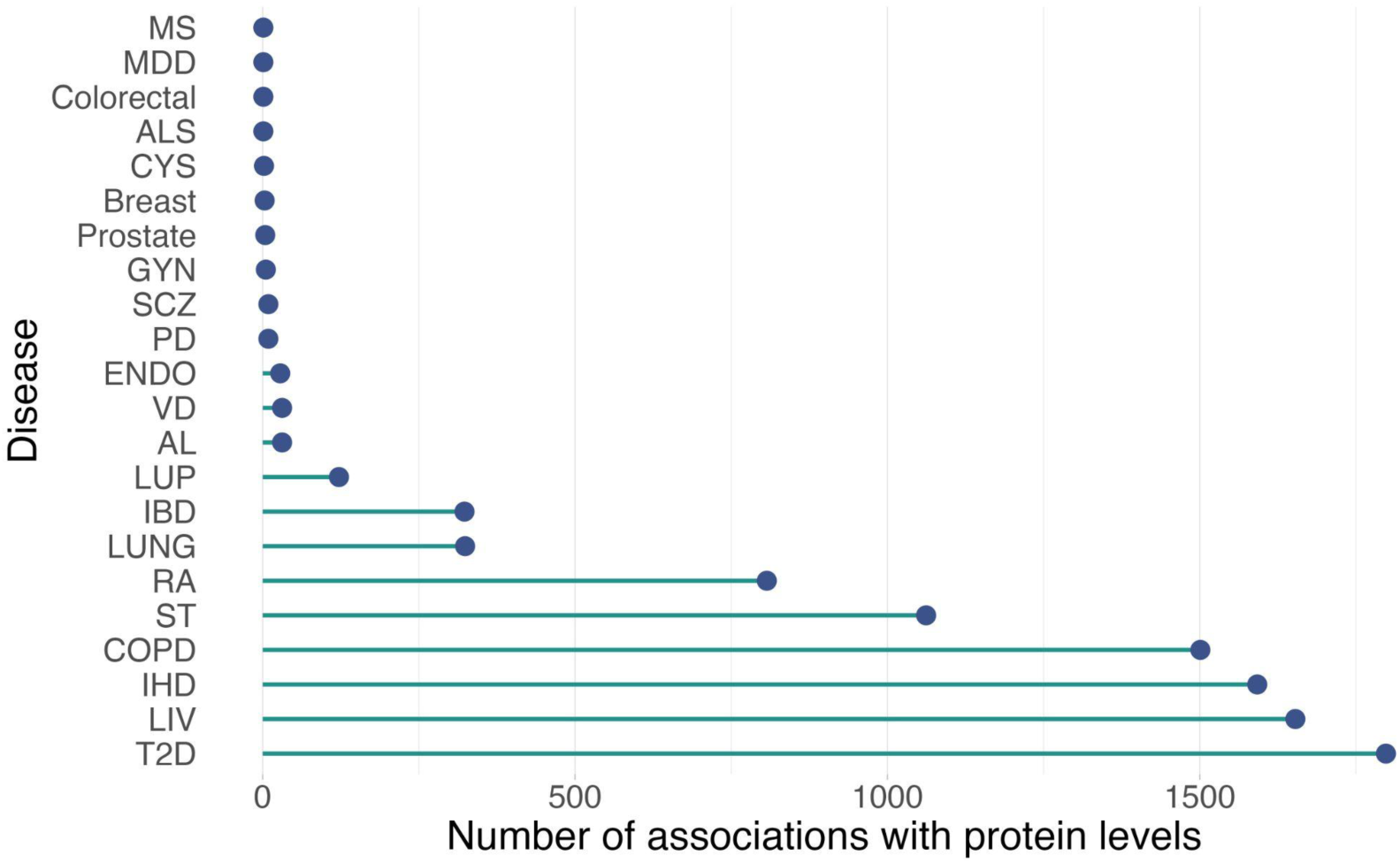
Number of associations between plasma proteins and 22 incident disease outcomes. 9,308 significant associations involving 2,122 proteins and 22 incident disease outcomes are shown (Bonferroni-adjusted *P*-value < 7.44 x 10^-7^). (MS = multiple sclerosis; MDD = major depressive disorder; Colorectal = colorectal cancer; ALS = amyotrophic lateral sclerosis; GYN = gynecological cancers; CYS = cystitis; Breast = breast cancer; Prostate = prostate cancer; PD = Parkinson’s disease; SCZ = schizophrenia; ENDO = endometriosis; AL = Alzheimer’s dementia; VD = vascular dementia; LUP = systemic lupus erythematosus; LUNG = lung cancer; IBD = inflammatory bowel disease; RA = rheumatoid arthritis; ST = ischemic stroke; COPD = chronic obstructive pulmonary disease; IHD = ischemic heart disease; LIV = Liver disease; T2D = Type 2 diabetes)

**Extended Data Fig. 2.**
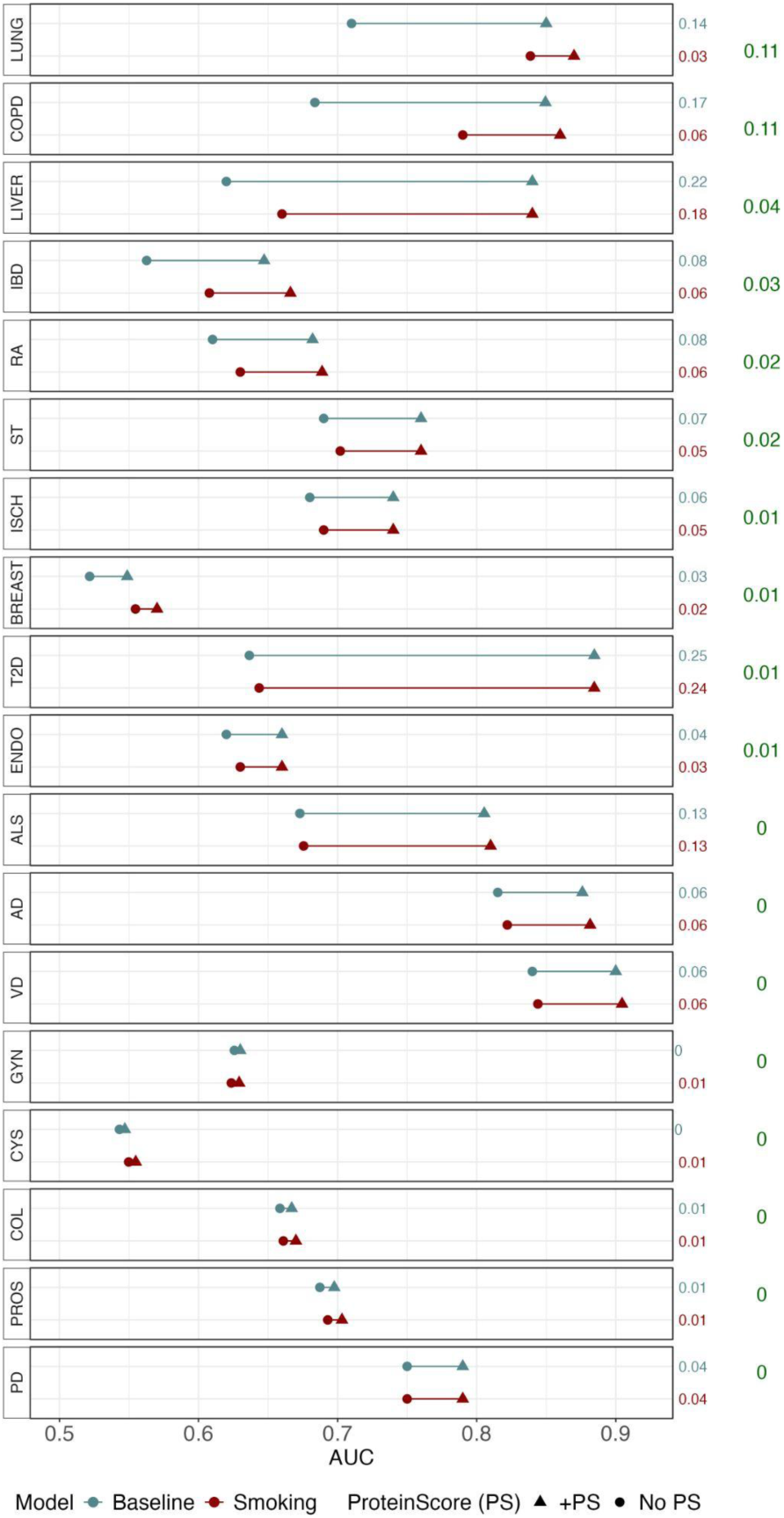
Performance of disease-based proteomic scores from Gadd et al.^7^. Differences in AUC between models with standard covariates and models with the addition of the disease ProteinScores (PS). Disease ProteinScores were developed and described in Gadd et al.^7^ In blue (baseline), models without PS consist of age and sex in which diseases were not sex-stratified. In red (smoking), models without PS consist of age, sex, and self-reported smoking status. To the right of the plot, the first column of numbers shows the differences in AUC with the addition of the PS per model; the second column shows Δ+PS in baseline minus Δ+PS in smoking.

**Extended Data Fig. 3.**
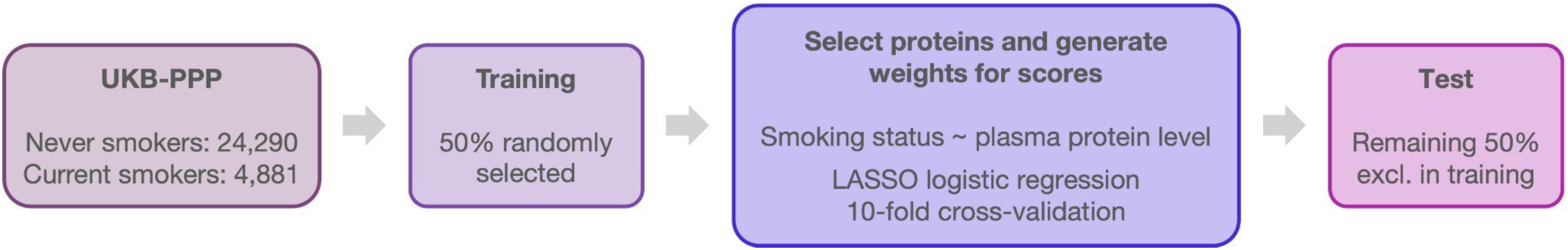
Summary of SmokingPS development. UKB-PPP participants who self-reported never and current smoking status were used for developing the SmokingPS. 50% were randomly assigned to the training group; the remaining 50% were assigned to the test group. Ratios of never to current smokers in each group were similar. LASSO regression with 10-fold cross validation was used to select proteins out of the 2,923 proteins measured and derive weighting coefficients for the selected proteins.

**Extended Data Fig. 4.**
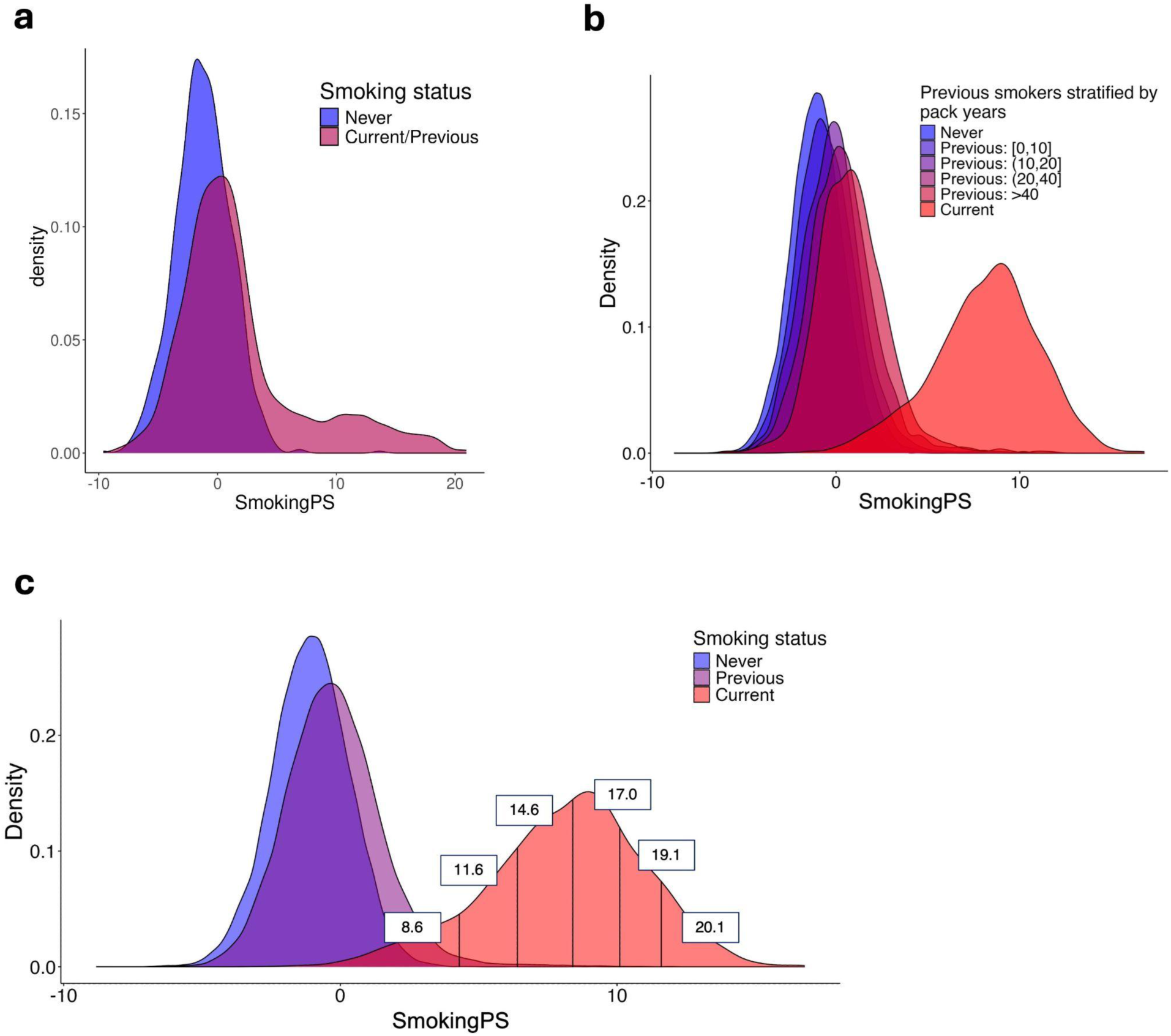
SmokingPS distributions in FinnGen and across previous and current smokers in UKB. **a**, Density plot of the SmokingPS in FinnGen participants (N = 1,862), stratified by self-reported never and current/previous smoking status. Current and previous smokers are grouped together due to gaps in timing between the collection of smoking information and proteomic sampling. **b,** Density plot of SmokingPS with self- reported previous smokers in UKB stratified by pack years, calculated as number of cigarettes smoked per day, divided by twenty, multiplied by number of years smoking. **c,** Density plot of SmokingPS with self-reported current smokers divided into bins of SmokingPS at 0.10, 0.25, 0.50, 0.75, and 0.90 quantiles. Cigarettes smoked per day were averaged across individuals in each bin; each average is reported on the density plot.

**Extended Data Fig. 5.**
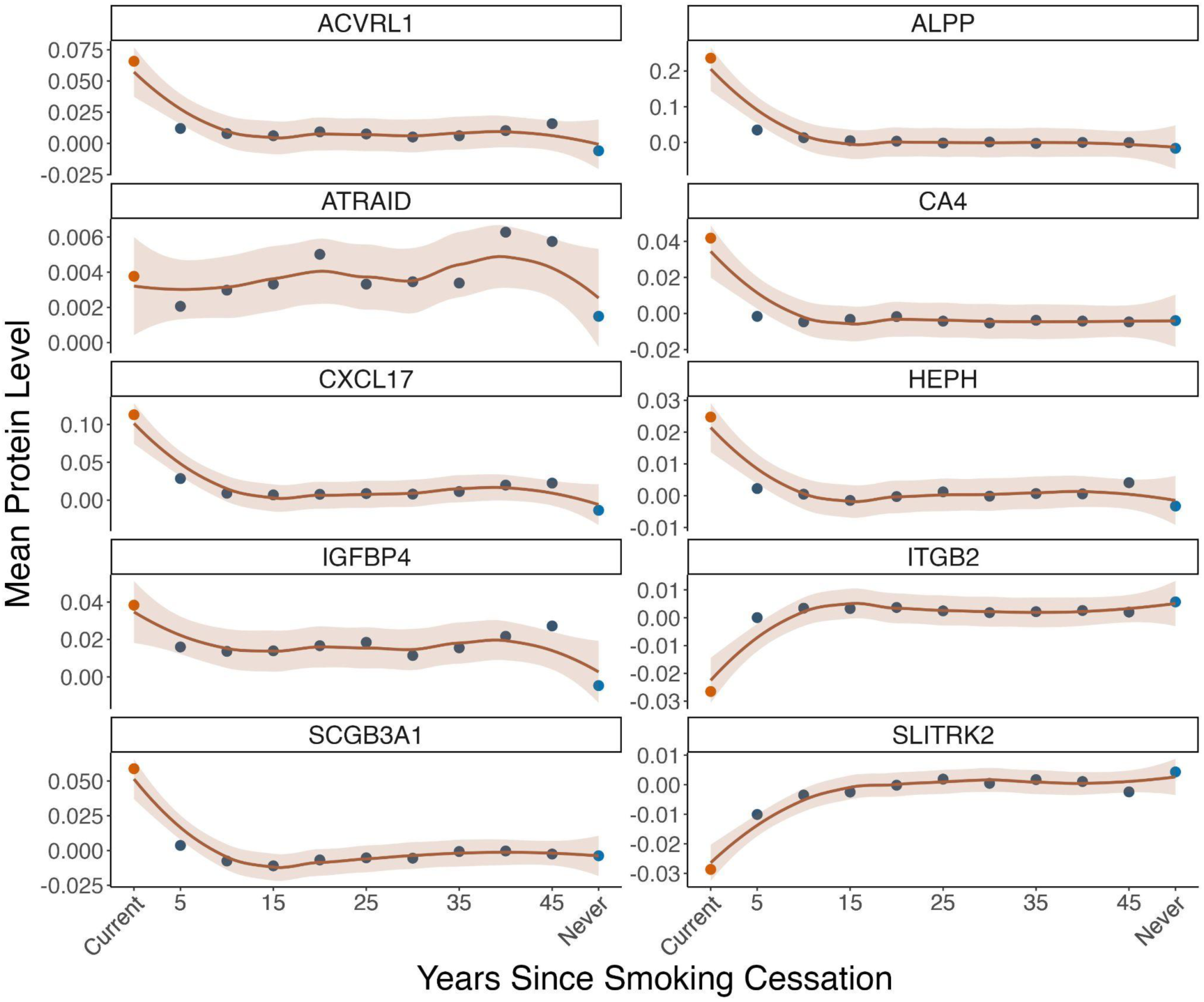
Average protein levels of top-weighted proteins in SmokingPS across groups of previous smokers. X-axis represents years since smoking cession, grouped in 5-year intervals for former smokers: (0,5], (10,15], etc. Average protein levels in current smokers and never smokers are included for reference.

**Extended Data Fig. 6.**
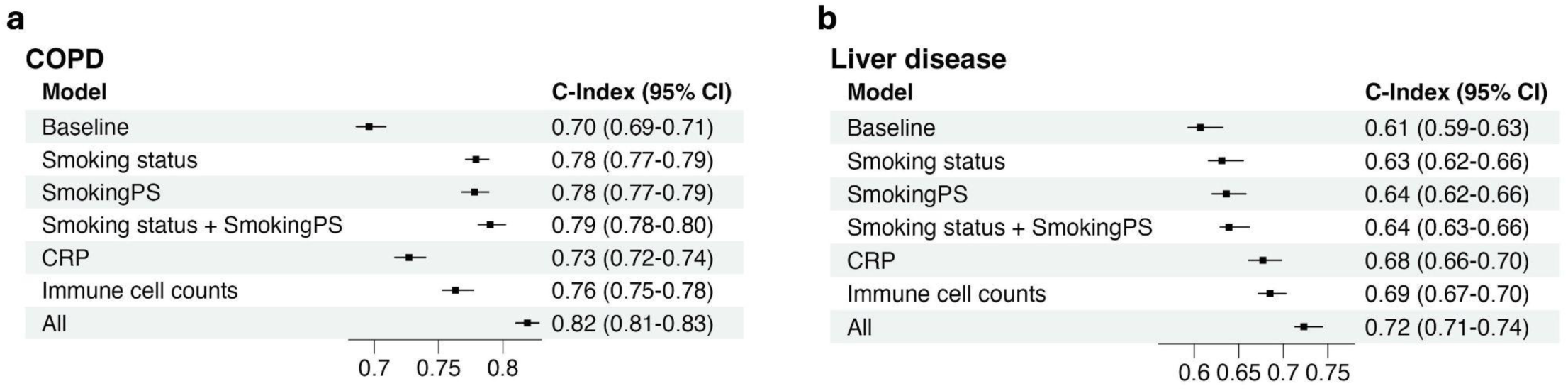
Comparison of SmokingPS and other models for prediction of incident COPD and liver disease. Associations between incident disease and models with various predictors, shown on *y*-axis, using Cox PH. Baseline model includes age, sex, age^2^, age × sex, age^2^ × sex, and first 10 genetic PCs. All models following baseline include these variables, as well as the predictor listed. “CRP” indicates C-reactive protein. “Immune cell counts” include neutrophil, eosinophil, basophil, monocyte, lymphocyte, and white blood cell counts. “All” indicates the baseline variables, smoking status, SmokingPS, CRP, and immune cell counts. C-index is shown on the *x-*axis and listed alongside 95% confidence intervals. **a,** Incident disease outcome is COPD (cases = 1,973, controls = 44,765). **b,** Incident disease outcome is liver disease (cases = 328, controls = 46,913).

**Extended Data Fig. 7.**
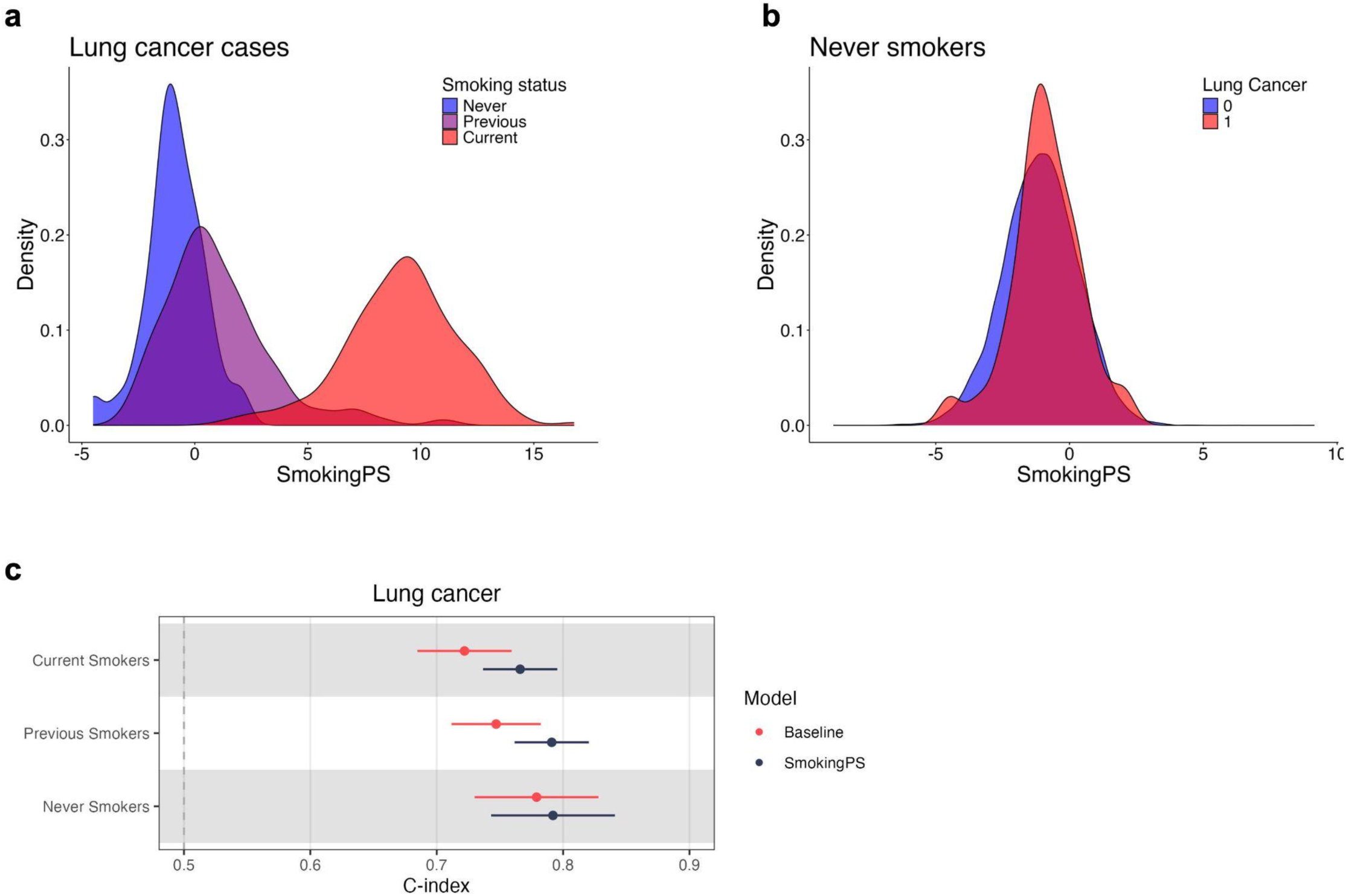
SmokingPS in individuals with lung cancer stratified by smoking status. **a,** Density plot of SmokingPS in individuals with lung cancer, stratified by self-reported smoking status. **b,** Density plot of SmokingPS in never smokers, stratified by incident lung cancer outcome. **c,** Cox PH associations between incident lung cancer and (1) baseline model of age, sex, age^2^, age × sex, age^2^ × sex, and first 10 genetic PCs and (2) baseline model with SmokingPS. Cox PH associations were conducted in individuals stratified by self- reported smoking status.

**Extended Data Fig. 8.**
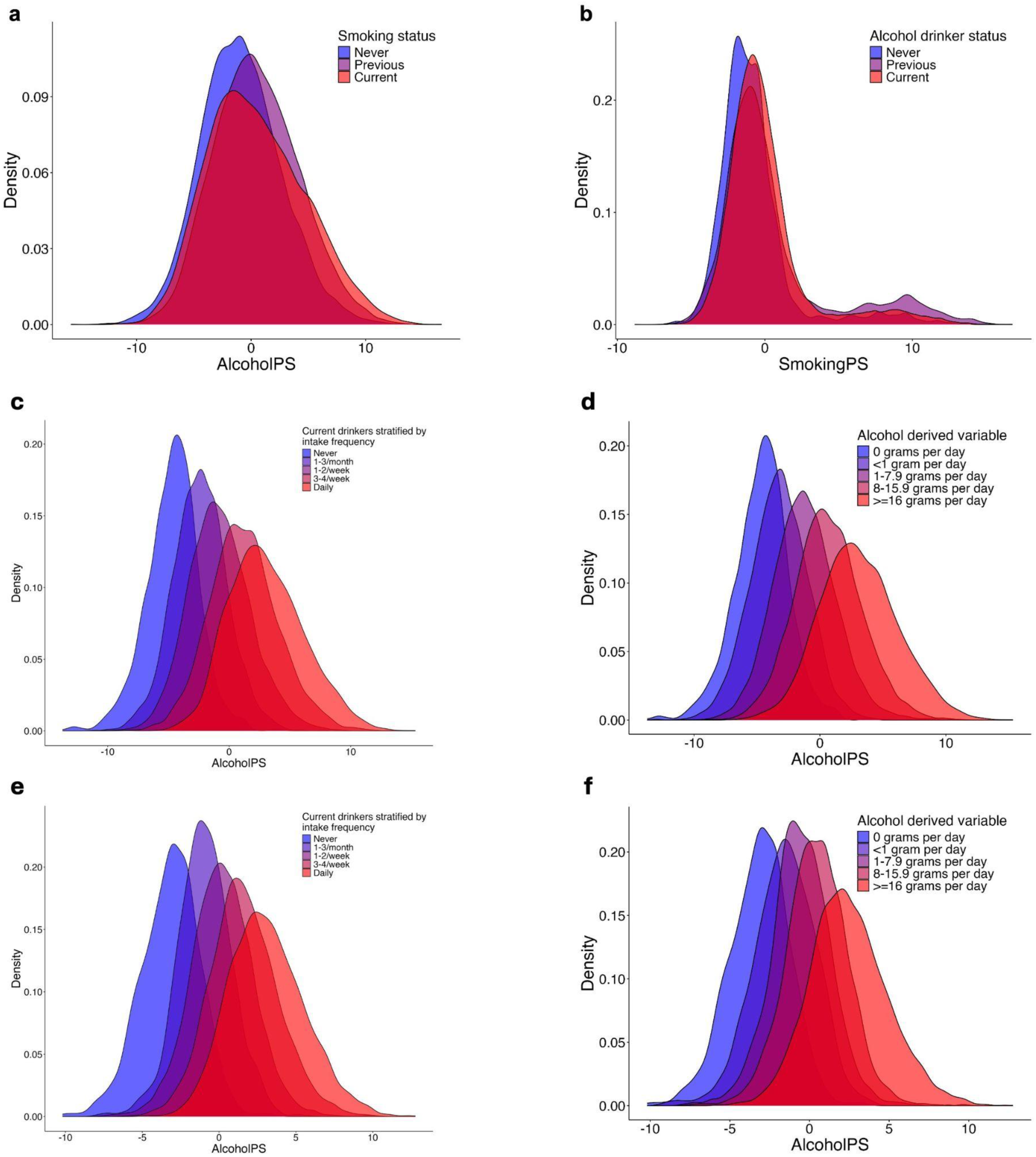
Comparisons of AlcoholPS and SmokingPS, and AlcoholPS trained in females and males separately. **a,** Density plot of AlcoholPS in individuals with self-reported smoking information, stratified by smoking status. **b,** Density plot of SmokingPS in individuals with self-reported alcohol intake information, stratified by alcohol drinker status. Density plot of AlcoholPS in current drinkers stratified by self-reported number of drinks per week or month in **c,** females and **e,** males. Self-reported never drinkers shown for reference. Density plot of AlcoholPS in current drinkers stratified by derived grams of alcohol intake per day in **d,** females and **f,** males.

**Extended Data Fig. 9.**
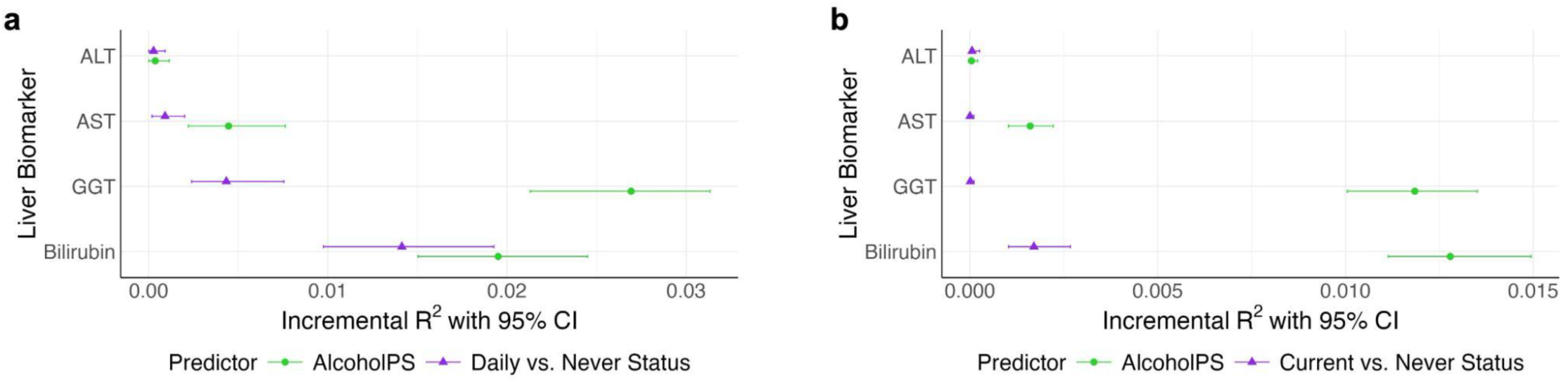
Associations between liver biomarkers and AlcoholPS. Comparisons of variance explained in liver biomarkers on *y-*axis by AlcoholPS and **a,** daily vs. never drinking status or **b,** current vs. never drinking status. Incremental *R^2^* was estimated as improvement in *R^2^* with inclusion of either AlcoholPS or alcohol intake status, comparing the two models: (1) baseline model (biomarker ∼ age + sex + age^2^ + age*sex + age^2^*sex) and (2) full model (biomarker ∼ AlcoholPS or alcohol intake status + age + sex + age^2^ + age*sex + age^2^*sex. (ALT = alanine aminotransferase, AST = aspartate aminotransferase, GGT = gamma-glutamyl transferase)

## References

1. Gudjonsson, A. et al. A genome-wide association study of serum proteins reveals shared loci with common diseases. Nat. Commun. 13, 480 (2022).

2. Emilsson, V. et al. Co-regulatory networks of human serum proteins link genetics to disease. Science 361, 769–773 (2018).

3. Sun, B. B. et al. Genomic atlas of the human plasma proteome. Nature 558, 73–79 (2018).

4. Ferkingstad, E. et al. Large-scale integration of the plasma proteome with genetics and disease. Nat. Genet. 53, 1712–1721 (2021).

5. Koprulu, M. et al. Proteogenomic links to human metabolic diseases. Nat. Metab. 5, 516–528 (2023).

6. Sun, B. B. et al. Plasma proteomic associations with genetics and health in the UK Biobank. Nature 622, 329–338 (2023).

7. Gadd, D. A. et al. Blood protein assessment of leading incident diseases and mortality in the UK Biobank. Nat Aging (2024) doi:10.1038/s43587-024-00655-7.

8. Carrasco-Zanini, J., et al. Proteomic prediction of common and rare diseases. bioRxiv (2023) doi:10.1101/2023.07.18.23292811.

9. Deng, Y.-T. et al. Atlas of the plasma proteome in health and disease in 53,026 adults. Cell 188, 253–271.e7 (2025).

10. Schuermans, A. et al. Integrative proteomic analyses across common cardiac diseases yield new mechanistic insights and enhanced prediction. medRxiv (2023) doi:10.1101/2023.12.19.23300218.

11. Ahsan, H. Monoplex and multiplex immunoassays: approval, advancements, and alternatives. Comp. Clin. Path. 31, 333–345 (2022).

12. Bowman, W. S. et al. Proteomic biomarkers of progressive fibrosing interstitial lung disease: a multicentre cohort analysis. Lancet Respir. Med. 10, 593–602 (2022).

13. Gudmundsdottir, V. et al. Circulating protein signatures and causal candidates for type 2 diabetes. Diabetes 69, 1843–1853 (2020).

14. Schuermans, A. et al. Genetic associations of circulating cardiovascular proteins with gestational hypertension and preeclampsia. JAMA Cardiol. 9, 209–220 (2024).

15. Chong, M. et al. Novel drug targets for ischemic stroke identified through Mendelian randomization analysis of the blood proteome. Circulation 140, 819–830 (2019).

16. Lu, T., Forgetta, V., Greenwood, C. M. T., Zhou, S. & Richards, J. B. Circulating proteins influencing psychiatric disease: A Mendelian randomization study. Biol. Psychiatry 93, 82–91 (2023).

17. Gong, W. et al. Genomics-driven integrative analysis highlights immune-related plasma proteins for psychiatric disorders. J. Affect. Disord. 370, 124–133 (2024).

18. Bourgault, J. et al. Proteome-wide Mendelian randomization identifies causal links between blood proteins and acute pancreatitis. Gastroenterology 164, 953–965.e3 (2023).

19. Zhang, S. et al. IDDF2024-ABS-0203 Causal links between plasma proteome and digestive system diseases: mendelian randomization analysis. in Basic Gastroenterology A143.1–A143 (BMJ Publishing Group Ltd and British Society of Gastroenterology, 2024).

20. Su, C.-Y. et al. Multi-ancestry proteome-phenome-wide Mendelian randomization offers a comprehensive protein-disease atlas and potential therapeutic targets. medRxiv (2024) doi:10.1101/2024.10.17.24315553.

21. Zhao, H. et al. Proteome-wide Mendelian randomization in global biobank meta-analysis reveals multi-ancestry drug targets for common diseases. Cell Genom 2, None (2022).

22. Guo, Y. et al. Plasma proteomic profiles predict future dementia in healthy adults. *Nat*. Aging 4, 247–260 (2024).

23. Chan, K. H. et al. An exposome-wide assessment of 6600 SomaScan proteins with non-genetic factors in Chinese adults. medRxiv (2024) doi:10.1101/2024.10.24.24316041.

24. Carrasco-Zanini, J. et al. Mapping biological influences on the human plasma proteome beyond the genome. Nat. Metab. 6, 2010–2023 (2024).

25. Isaac, S. et al. Human plasma proteomics links modifiable lifestyle exposome to disease risk. medRxiv 2025.05.07.25327178 (2025) doi:10.1101/2025.05.07.25327178.

26. Burgess, S. & Thompson, S. G. Interpreting findings from Mendelian randomization using the MR-Egger method. Eur. J. Epidemiol. 32, 377–389 (2017).

27. Bergeron, N., Phan, B. A. P., Ding, Y., Fong, A. & Krauss, R. M. Proprotein convertase subtilisin/Kexin type 9 inhibition: A new therapeutic mechanism for reducing cardiovascular disease risk. Circulation 132, 1648–1666 (2015).

28. Raulin, A.-C. et al. ApoE in Alzheimer’s disease: pathophysiology and therapeutic strategies. Mol. Neurodegener. 17, 72 (2022).

29. Penrose, H. M. et al. Ulcerative colitis immune cell landscapes and differentially expressed gene signatures determine novel regulators and predict clinical response to biologic therapy. Sci. Rep. 11, 9010 (2021).

30. Feiner, J. et al. 221-LB: Identification of PAM as novel monogenic diabetes gene. Diabetes 72, 221–LB (2023).

31. Sun, Z. et al. Comprehensive mendelian randomization analysis of plasma proteomics to identify new therapeutic targets for the treatment of coronary heart disease and myocardial infarction. J. Transl. Med. 22, 404 (2024).

32. Jin, H. et al. Smoking, COPD, and 3-nitrotyrosine levels of plasma proteins. Environ. Health Perspect. 119, 1314–1320 (2011).

33. Elisia, I. et al. The effect of smoking on chronic inflammation, immune function and blood cell composition. Sci. Rep. 10, 19480 (2020).

34. Chan, K. H. et al. Tobacco smoking and risks of more than 470 diseases in China: a prospective cohort study. Lancet Public Health 7, e1014–e1026 (2022).

35. Zhang, J.-C. et al. TGF-β/BAMBI pathway dysfunction contributes to peripheral Th17/Treg imbalance in chronic obstructive pulmonary disease. Sci. Rep. 6, 31911 (2016).

36. Kurki, M. I. et al. FinnGen provides genetic insights from a well-phenotyped isolated population. Nature 613, 508–518 (2023).

37. Saint-André, V. et al. Smoking changes adaptive immunity with persistent effects. Nature 626, 827–835 (2024).

38. Chybowska, A. D. et al. A blood- and brain-based EWAS of smoking. Nat. Commun. 16, 3210 (2025).

39. Joehanes, R. et al. Epigenetic signatures of cigarette smoking. Circ. Cardiovasc. Genet. 9, 436–447 (2016).

40. McCartney, D. L. et al. Epigenetic signatures of starting and stopping smoking. EBioMedicine 37, 214–220 (2018).

41. Fang, F., Andersen, A. M., Philibert, R. & Hancock, D. B. Epigenetic biomarkers for smoking cessation. Addict. Neurosci. 6, 100079 (2023).

42. Frickmann, H. et al. The influence of virus infections on the course of COPD. Eur. J. Microbiol. Immunol. (Bp.) 2, 176–185 (2012).

43. Mortimer, K. et al. Household air pollution and COPD: cause and effect or confounding by other aspects of poverty? Int. J. Tuberc. Lung Dis. 26, 206–216 (2022).

44. Xue, A. et al. Genome-wide analyses of behavioural traits are subject to bias by misreports and longitudinal changes. Nat. Commun. 12, 20211 (2021).

45. Hou, K. et al. Causal effects on complex traits are similar for common variants across segments of different continental ancestries within admixed individuals. Nat. Genet. 55, 549–558 (2023).

46. Hu, S. et al. Fine-scale population structure and widespread conservation of genetic effect sizes between human groups across traits. Nat. Genet. 57, 379–389 (2025).

47. Bycroft, C. et al. The UK Biobank resource with deep phenotyping and genomic data. Nature 562, 203– 209 (2018).

48. Argentieri, M. A. et al. Proteomic aging clock predicts mortality and risk of common age-related diseases in diverse populations. Nat. Med. 30, 2450–2460 (2024).

49. Choi, K. W. et al. An exposure-wide and Mendelian randomization approach to identifying modifiable factors for the prevention of depression. Am. J. Psychiatry 177, 944–954 (2020).

50. Evangelou, E. et al. New alcohol-related genes suggest shared genetic mechanisms with neuropsychiatric disorders. *Nat*. Hum. Behav. 3, 950–961 (2019).

51. Therneau, T. A package for survival analysis in R. Preprint at https://cran.r-project.org/web/packages/survival/vignettes/survival.pdf (2024).

52. Okada, Y. et al. Genetics of rheumatoid arthritis contributes to biology and drug discovery. Nature 506, 376–381 (2014).

53. Willer, C. J., Li, Y. & Abecasis, G. R. METAL: fast and efficient meta-analysis of genomewide association scans. Bioinformatics 26, 2190–2191 (2010).

54. Chen, S. et al. A genomic mutational constraint map using variation in 76,156 human genomes. Nature 625, 92–100 (2024).

55. Hemani, G. et al. The MR-Base platform supports systematic causal inference across the human phenome. Elife 7, (2018).

56. Burgess, S. & Yavorska, O. MendelianRandomization: Mendelian randomization package. CRAN: Contributed Packages The R Foundation 10.32614/cran.package.mendelianrandomization (2016).

57. Enea, M. _speedglm: Fitting Linear and Generalized Linear Models to Large Data Sets_. (2023).

58. Gu, Z., Eils, R. & Schlesner, M. Complex heatmaps reveal patterns and correlations in multidimensional genomic data. Bioinformatics 32, 2847–2849 (2016).

59. Friedman, J., Hastie, T. & Tibshirani, R. Regularization paths for generalized linear models via coordinate descent. J. Stat. Softw. 33, 1–22 (2010).

60. Robin, X. et al. pROC: an open-source package for R and S+ to analyze and compare ROC curves. BMC Bioinformatics 12, 77 (2011).

61. Karczewski, K. J. et al. Pan-UK Biobank GWAS improves discovery, analysis of genetic architecture, and resolution into ancestry-enriched effects. (2024) doi:10.1101/2024.03.13.24303864.

62. Dayimu, A. _forestploter: Create a Flexible Forest Plot_. R package version 1.1.2 https://CRAN.R-project.org/package=forestploter (2024).

